# The evolving epidemiology of scrub typhus in Thailand (2003–2024): insights from latent process modelling of national surveillance data

**DOI:** 10.64898/2026.04.20.26351270

**Authors:** Phrutsamon Wongnak, Kittipong Chaisiri, Carlo Perrone, Karine Chalvet-Monfray, Darin Areechokchai, Wirichada Pan-ngum

**Affiliations:** Mahidol-Oxford Tropical Medicine Research Unit (MORU), Faculty of Tropical Medicine, Mahidol University, Bangkok, 10400 Thailand; Department of Helminthology, Faculty of Tropical Medicine, Mahidol University, Bangkok, 10400 Thailand; Centre for Tropical Medicine and Global Health, Nuffield Department of Medicine, University of Oxford, Oxford OX3 7BN, United Kingdom; Université de Lyon, UMR EPIA, INRAE, VetAgro Sup, 69280 Marcy l’Etoile, France; Division of Vector Borne Diseases, Department of Disease Control, Ministry of Public Health, Nonthaburi, 11000 Thailand; Department of Tropical Hygiene, Faculty of Tropical Medicine, Mahidol University, Bangkok, 10400 Thailand

**Keywords:** Passive surveillance, Underreporting, *Orientia tsutsugamushi*, Seasonality, Thailand

## Abstract

**Background:** Scrub typhus is a major yet neglected vector-borne disease in Thailand, where it has been nationally notifiable for over two decades. However, long-term changes in its epidemiology, including reporting proportions, transmission intensity, disease severity, and seasonal patterns, have not been comprehensively characterised at the national level.

**Methodology:** We analysed 22 years of national surveillance data for scrub typhus in Thailand (2003–2024) using a latent process model that jointly fits reported cases with published nationwide seroprevalence data and antibody kinetics to estimate reporting proportions and underlying transmission dynamics across all 77 provinces of Thailand.

**Findings:** Over the 22-year study period, 143096 cases and 119 deaths were reported nationally. Estimated reporting proportion broadly mirrored transmission intensity, being higher in high-burden regions and lower elsewhere. Estimated reporting proportion declined synchronously across all regions during the COVID-19 pandemic, followed by rapid rebound by 2024. After adjusting for estimated reporting, the estimated force of infection was highest in the northern provinces but also substantial in the northeast and south, with upward trends in some provinces. Susceptibility among older adults aged 65 and above increased progressively over the study period, reversing the pattern observed two decades earlier. Case-fatality among reported cases in the 25–35-year reference group was low and declined from 0.15% (95% Credible Interval [CrI]: 0.07– 0.30%) to 0.06% (95% CrI: 0.02–0.12%), while relative case-fatality among adults above 65 remained elevated across all periods. Three seasonal patterns, differing mainly in peak timing, were identified and remained largely stable over time.

**Conclusion:** Over two decades, scrub typhus transmission in Thailand appears to extend well beyond its traditionally recognised northern focus, with estimated substantial burden in regions that have received less attention, while the demographic profile of those most affected has shifted progressively toward older adults. These findings support the need for regionally tailored surveillance, age-targeted clinical preparedness, and sustained investment in understanding the ecological drivers of transmission.

**Author summary:** Scrub typhus is a common but neglected cause of fever in Thailand, where it has been reported through the national surveillance system for over two decades. However, trends in reported cases can be misleading because they reflect not only true changes in transmission but also variation in diagnosis and reporting over time and across regions. We developed a model that combines surveillance data with seroprevalence surveys and antibody kinetics to estimate transmission and reporting jointly, allowing us to estimate how transmission intensity, disease severity, and seasonal patterns have evolved from 2003 to 2024 across all 77 provinces. Our estimates suggest that substantial transmission occurs not only in the well-studied northern provinces but also in the northeast and south, where the disease has received less attention. Susceptibility has progressively shifted toward older adults, who also face elevated case-fatality, while three seasonal patterns vary by region and have remained largely stable over time. These findings suggest that scrub typhus control in Thailand requires a shift from a predominantly northern focus toward regionally tailored strategies that account for local transmission timing, an ageing at-risk population, and the ecological drivers that sustain transmission in each setting.

## Introduction

Scrub typhus remains a neglected vector-borne disease in Thailand, more than seven decades after its first report in the country in 1952^1^. The disease is caused by obligate intracellular bacteria of the genus *Orientia* and is transmitted to humans through the bite of infected larval trombiculid mites (Acari: Trombiculidae), commonly known as chiggers. Infection can lead to severe febrile illness with systemic complications and, if not treated promptly, may result in substantial morbidity and death^2^. Although effective antibiotics are available, misdiagnosis^3,4^ and uneven access to care^5^ continue to cause preventable deaths from scrub typhus.

Understanding how the epidemiology of scrub typhus evolves over the long term is important for assessing disease burden, guiding public health interventions, and identifying emerging trends. This is particularly relevant for scrub typhus, as the ecology and seasonal activity of its chigger vectors are closely tied to temperature, rainfall, and humidity^6^, while human exposure is shaped by land use, agricultural practices, and demographic shifts. In Thailand, rapid urbanisation, an ageing agricultural workforce, and ongoing environmental change may all be reshaping the transmission landscape, making sustained epidemiological monitoring essential.

Thailand’s nationwide passive surveillance system for scrub typhus, operated through the National Disease Surveillance database (R506), provides a valuable resource for such monitoring. Government and private healthcare facilities across all 77 provinces report clinically suspected and laboratory-confirmed cases, yielding a continuous record of notifications spanning more than two decades. Using these data from 2003 to 2018, Wangrangsimakul et al.^7^ described rising incidence, geographic concentration in the northern region, and associations with agricultural occupation and environmental factors in Chiang Rai Province.

Passive surveillance data, however, do not directly reflect underlying transmission. Reported case counts are shaped by healthcare-seeking behaviour, diagnostic capacity, clinical awareness, and reporting practices, all of which may vary across regions and over time. Temporal trends in such data therefore represent a mixture of true epidemiological change and variation in detection and reporting and may be misinterpreted if these processes are not explicitly accounted for. Several approaches have been proposed to address underreporting and under-ascertainment in infectious disease surveillance, including capture-recapture methods, multiplier approaches, and model-based estimation^8^, each with different data requirements and assumptions.

In this study, we extend the analysis of Wangrangsimakul et al.^7^ through 2024 and address the limitations of passive surveillance by jointly modelling reported case data with nationwide seroprevalence data and published antibody kinetics. This approach allows us to estimate temporal changes in reporting and underlying transmission jointly, and to examine long-term shifts over 22 years across four key dimensions: reporting proportions, transmission intensity, disease severity, and seasonal patterns. Our aim is to characterise how the epidemiology of scrub typhus in Thailand has evolved over this period and to identify priorities for surveillance, research, and public health action.

## Materials and methods

### Data

#### Disease Surveillance data

Scrub typhus reported cases and deaths from 2003 to 2024 were obtained from the National Disease Surveillance (Report 506) database^9^, publicly available passive disease surveillance system by the Division of Epidemiology, Department of Disease Control, Ministry of Public Health (MoPH), Thailand. Reported cases in the database included suspected, probable, and confirmed cases, defined as follows : (1) suspected cases fulfilled the clinical criteria and had a history of environmental exposure; (2) probable cases fulfilled the clinical criteria, had a history of contact with confirmed cases, and met the presumptive laboratory diagnosis criteria, that is an immunochromatographic assay; and (3) confirmed cases fulfilled the clinical criteria and met the laboratory diagnosis criteria, including polymerase chain reaction, indirect immunofluorescent assay, indirect immunoperoxidase test, or Weil Felix test to OX K^10^.

The annual disease surveillance data consist of two sub-datasets describing the epidemiology for the same year: (1) monthly reported scrub typhus cases and deaths; and (2) annual reported scrub typhus cases and deaths by age group. Data are reported by province, covering all 77 provinces of Thailand across all 13 health regions.

#### Seroprevalence data

Data on the proportion of scrub typhus sero-positivity were extracted from Gonwong et al., (2022)^11^ based on immunoglobulin G (IgG) measurements using ELISA among young men (average age 21 years at the time of measurement) across all provinces of Thailand in 2007-2008 and 2012. In this study, sero-positivity was defined using a cut-off of two standard deviations above the mean optical density of negative control sera. Reported seroprevalence was categorised into the following ranges: 10%, 11-15%, ≤16-20%, 21-25%, and 26-35%.

#### Population statistics data

Historical annual population sizes by province and age group were obtained from the Administrative Information System of the Bureau of Registration Administration, Ministry of Interior, Thailand^12^. Population data from January of each year were used. Average annual reported incidence and mortality were calculated as reported cases or deaths divided by person-years (the sum of annual population sizes), per 100,000 and 1,000,000 population, respectively, and were directly age-standardised to the pooled national population (2004–2024) as a sensitivity check.

### Ethics statement

Ethical approval was not required as this study relied solely on publicly available, de-identified secondary data with no individual-level information.

### Definitions of epidemiological quantities

Key quantities used throughout this study are defined in Table 1. Apart from reported cases, reported deaths and population size, all quantities are model-derived estimates rather than direct observations.

**Table 1.** Definitions of key epidemiological quantities.

| Quantity | Definition | Source |
|---|---|---|
| Reported case | Scrub typhus notification in the national surveillance system, comprising suspected, probable and confirmed cases | Data |
| Reported death | Death among reported cases notified to the surveillance system | Data |
| At-risk population | Resident population of a province or age group; existing immunity is absorbed into the force of infection (Eq 1) | Data |
| Clinical infection | Symptomatic infection, assumed to occur in a fixed proportion (clinical fraction, ν) of all infections; the remainder are subclinical | Model-derived |
| Reporting proportion (π) | Probability that a clinical infection is captured by the surveillance system as a reported case | Model-derived |
| Annual force of infection (FOI) | Annual hazard of infection per at-risk person in the 25–35-year reference age group, calculated as twelve times the baseline monthly force of infection (λ<sup>base</sup>) | Model-derived |
| Estimated incidence | Infections, clinical and subclinical, per 100,000 at-risk population per year, calculated as (1 − exp(−annual FOI)) × 100,000; not the incidence of reported cases | Model-derived |
| Relative susceptibility (ρ) | Force of infection in an age group relative to the 25–35-year reference group | Model-derived |
| Case-fatality rate (CFR) | Proportion of reported cases that result in a reported death; not the infection-fatality risk among all infections | Model-derived |
| Relative case-fatality rate (η) | Case-fatality rate in an age group relative to the 25–35-year reference group | Model-derived |

### Model descriptions

We formulated a Bayesian latent process model to jointly analyse monthly reported scrub typhus cases and deaths, age distributions, and province-level seroprevalence data across Thailand. The model operates at the monthly and provincial level, decomposing observed surveillance data into underlying epidemiological and operational processes.

#### Expected clinical cases and deaths

The latent number of expected clinical cases at month *t*, age group *a*, in province *s*, denoted as *I*_*t,a,s*_ modelled as a function of the force of infection acting on the population as follows:

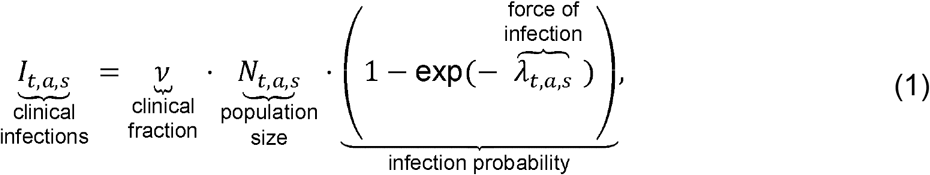

where *v* is the clinical fraction representing the proportion of infections that develop clinical disease, *N*_*t,a,s*_ is the population size, and *λ*_*t,a,s*_ is the effective force of infection for individuals in age group *a* in province *s*, integrated over month *t*. The term 1 – exp(–*λ*_*t,a,s*_) therefore represents the probability of infection over the monthly time step, derived from the assumption that infections follow a Poisson process. The formulation represented in Eq (1) does not imply that the entire population is susceptible to the infection. Instead, partial immunity, waning protection, and strain turnover are implicitly absorbed into *λ*_*t,a,s*_ and it should therefore be interpreted as conditional on the existing immune landscape. In the absence of precise estimates for the Thai population, the clinical fraction *v* was fixed at 0.15 for all age groups, based on the ratio of clinical scrub typhus cases to seroconversion incidence (1238/8123) reported in a recent cohort study in rural India^13^.

The expected number of reported clinical cases, denoted as *C*_*t*_,_*a*_,_*s*_, is modelled by accounting for imperfect surveillance through year- and region-specific reporting proportion *π*_*y*(*t*)_,_*r*(*s*)_ ∈, which represents the probability that a true clinical infection is reported by the surveillance system. Since individual provinces (indexed by *s*) may report too few cases to reliably estimate their own reporting proportions, provinces are grouped following the 13 MoPH Health Regions (Fig. 1C), and all provinces within the same region share a single reporting proportion:

**Figure 1.**
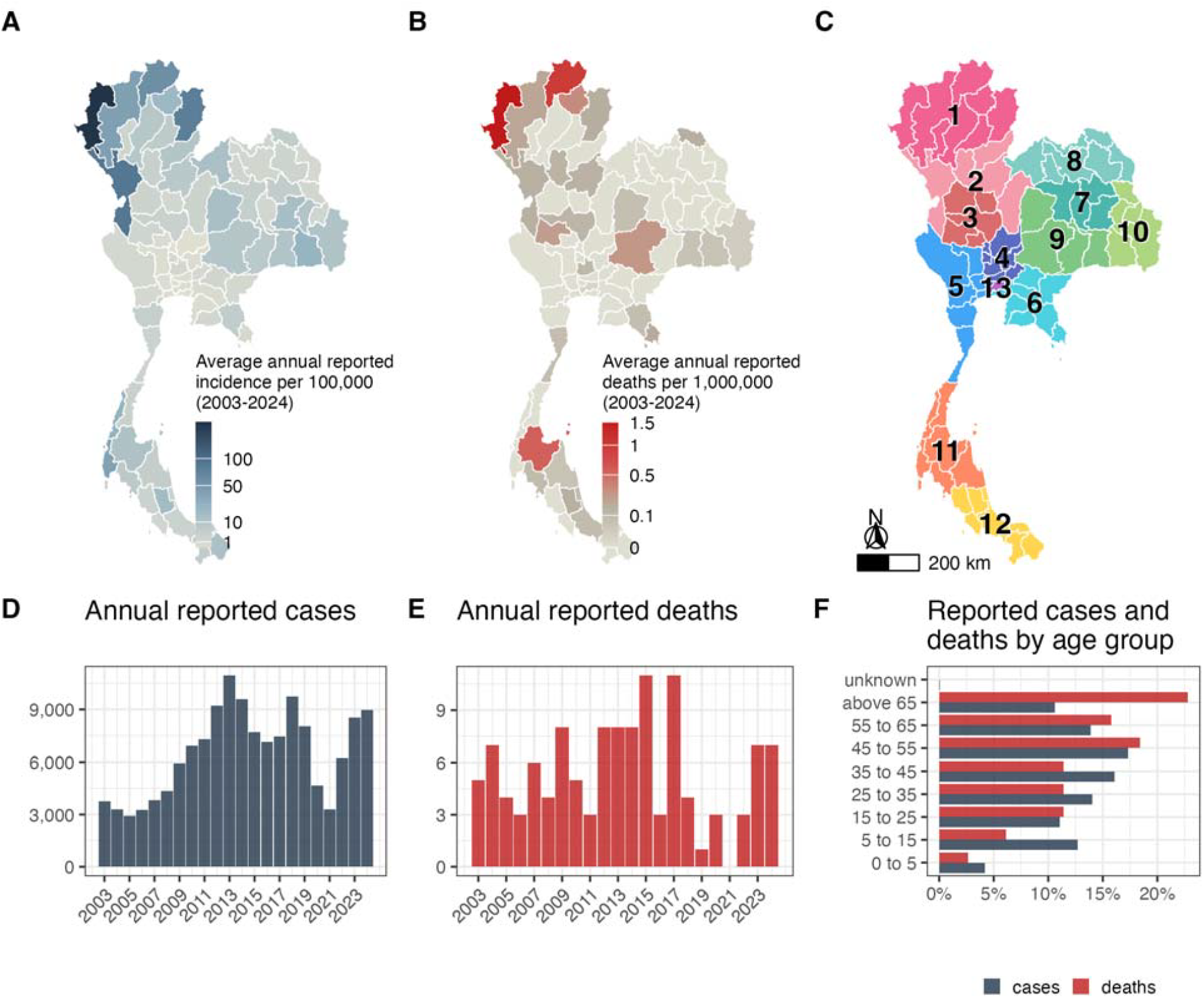
Reported scrub typhus cases and deaths in Thailand, 2003 to 2024. Reported cases comprise suspected, probable and confirmed cases notified to the national surveillance system. A: Average annual reported incidence per 100,000 population. B: Average annual reported deaths per 1,000,000 population. C: Health regions of Thailand. D: Trend in annual reported cases. E: Trend in annual reported deaths. F: Age distribution of reported cases and deaths over 2003 to 2024.

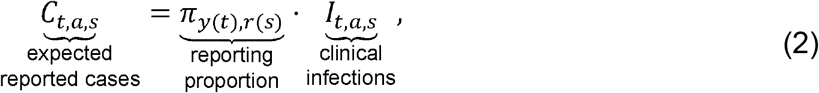

The reporting proportion is allowed to evolve over time following a region-specific random walk process (Supplementary material Text S1.1), capturing gradual changes in surveillance performance. In addition, although surveillance relies partly on imperfect diagnostic tests and some false positive reports are therefore expected, these are not modelled explicitly and are instead absorbed into the reporting process and accommodated through the overdispersed observation model. Similarly, the aggregated surveillance data did not distinguish suspected, probable and confirmed cases or record the diagnostic test used, so changes in diagnostic practice over time are also absorbed into the region-specific reporting proportion. Finally, the expected number of deaths is modelled as a function of reported cases:

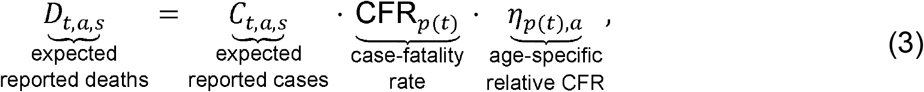

where *CFR*_*p*(*t*)_ is the baseline case-fatality rate during period *p*(*t*), and *η*_*p(t)*_,_*a*_ represents the relative case-fatality rate for age group *a* within period *p*(*t*). Periods were defined as blocks of 5-6 consecutive years. The 25–35-year age group was used as the reference category *a*\* such that *η*_*p*_,_*a*_* = 1.

### Force of infection

The force of infection *λ*_*t*_,_*a*_,_*s*_ is expressed as a multiplicative combination of baseline, seasonal, and age-specific components:

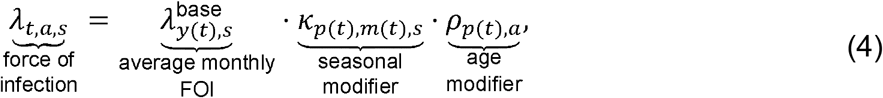

where 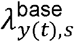denotes the average monthly (baseline) force of infection in year *y*(*t*) and province *s*. The baseline force of infection is allowed to evolve over years following a province-specific random walk process (Supplementary material Text S1.2), capturing gradual long-term changes in transmission intensity. The terms *κ*_*p*(*t*),*m*(*t*),*s*_ ∈ (0, ∞) is a seasonal modifier capturing within-year variation in transmission for province *s* defined within period *p*(*t*). Seasonality is modelled using a Fourier series with three harmonics (Supplementary material Text S1.3), allowing flexible but smooth seasonal patterns within each period. The number of harmonics was set to three based on empirical evaluation through posterior predictive checks, which confirmed that three harmonics provided adequate fit to the observed seasonal patterns without overfitting. Finally, *ρ*_*p*(*t*),*a*_ represents the relative susceptibility of age group *a* during period *p*(*t*). The 25-35-year age group was also used as the reference category *a*\* (*ρ*_*p,a*\*_=1).

### Seroprevalence

To incorporate seroprevalence data, we relate province-level serological measurements to the cumulative force of infection over time. Specifically, seroprevalence in province *s* in year *y*, denoted *p*_*y,s*_ is modelled as:

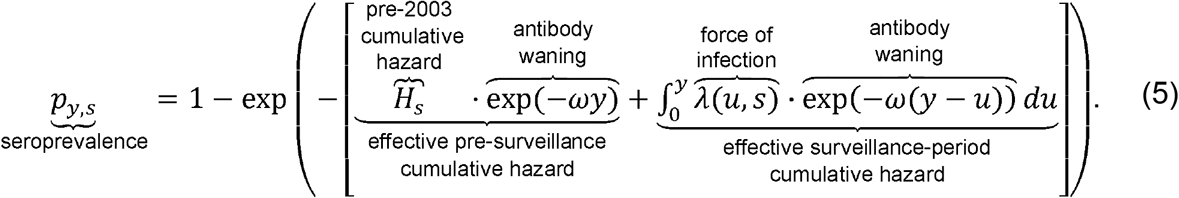

This formulation represents the accumulation of infection risk over time under a Poisson infection process, while allowing previously infected individuals to lose detectable IgG at a waning rate *ω*. Here, *u* is a dummy variable of integration representing time, ranging from the beginning of the surveillance period (*u* = 0) to the current year (*u* = *y*), and the term exp(–*ω*(*y* – *u*)) accounts for the decrease of the antibody over the interval (*y* – *u*) elapsed since infection at time *u*. The IgG waning rate *ω* was fixed at 0.54 year^−1^, corresponding to an average duration of 1.86 years (22.3 months) during which IgG levels remain above the seropositivity cut-off^14^. The term *H*_*s*_ represents a province-specific initial cumulative hazard, capturing pre-existing immunity at the start of surveillance, that is, infections occurring before 2003. The effective force of infection *λ*(*u,s*) is derived from the estimated baseline and seasonal transmission components. In practice, the model is implemented in discrete time with monthly steps, and the integral is approximated by a finite sum of past annual forces of infection:

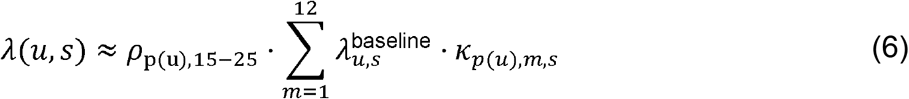

For each province, we define a province-level model-predicted seroprevalence *p*_*s*_, to be used in subsequent observation models, as the average of *p*_*t,s*_ across the years in which serological surveys were conducted (2007, 2008, and 2012). Because the serological surveys were conducted among young men (average age 21 years), model-predicted seroprevalence was evaluated for the 15–25-year age group, with the force of infection in Eqs (5) and (6) scaled by the relative susceptibility of this age group, *ρ*_*p*(*u*),15–25_. The serological data therefore inform transmission in the age group they represent, whereas transmission in other age groups is informed by the age distribution of reported cases through *ρ*_*p,a*_.

### Observation models

We link the monthly age-specific expected case reports *C*_*t,a,s*_ derived from the model to two marginalised surveillance datasets: monthly totals aggregated over age, 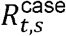, and annual age distributions aggregated over months, 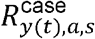.

Monthly totals are modelled using a negative binomial likelihood,

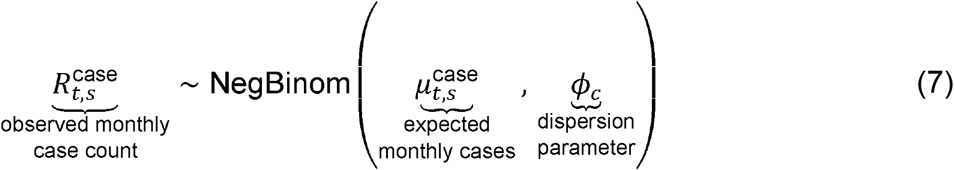

where, 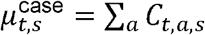, and *ϕ*_*c*_ represents a dispersion parameter for the negative binomial distribution. While annual age distributions are modelled using a multinomial likelihood conditional on the annual totals,

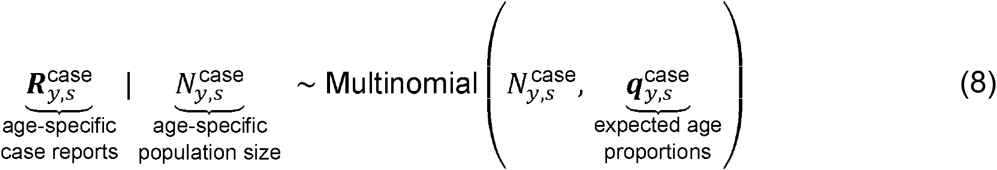

where 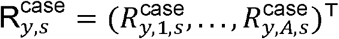 is the vector of observed age-specific case counts, and 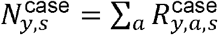 is the observed total number of reported cases in year *y* and province *s*. The vector 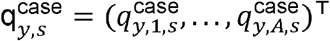 denotes the expected age proportions, defined as:

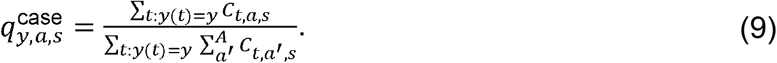

The likelihood of deaths is specified analogously, linking the expected deaths *D* _*t,a,s*_ from Eq. (3) to observed monthly totals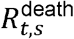 and annual age distributions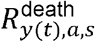. Years with incomplete or missing age information contribute only through the monthly total likelihood and are excluded from the multinomial likelihood.

In addition, seroprevalence data^11^ are incorporated using an interval-censored likelihood. Let 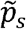 denote the latent unobserved realised seroprevalence measurement in province *s*. We assume that 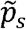 is normally distributed around model-predicted *p*_*s*_ with standard deviation *σ*_sero_. Since seroprevalence is only observed through reported bounds [*L*_*s*_, *U*_*s*_], the likelihood contribution is given by the probability that 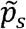 lies within this interval as:

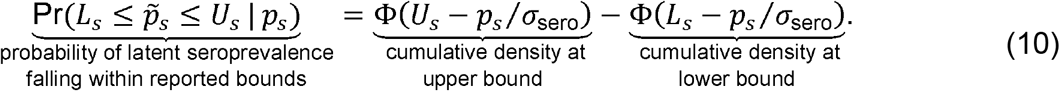

where *ϕ*(·) denotes the standard normal cumulative density function. We fix *σ*_sero_ = 0.01 so that the likelihood strongly penalises model predictions falling outside the reported interval, effectively treating the seroprevalence bounds as informative constraints.

### Parameter estimation

Model parameters were estimated within a hierarchical Bayesian framework using Hamiltonian Monte Carlo (HMC) sampling, implemented in Stan via the *rstan* package. We assigned weakly informative priors to all parameters to provide broad regularization while allowing the observed data to dominate the posterior distributions (Supplementary material Table S2). We ran four independent Markov Chain Monte Carlo (MCMC) chains of 2,000 iterations each, including a 1,000-iteration warm-up period. Post-warmup, chains were thinned every 4 samples, yielding a total of 1,000 posterior draws. Convergence and model fit were assessed using trace plots and posterior predictive checks to ensure the model adequately captured the observed temporal trends in scrub typhus reports for each province. To assess the influence of the fixed clinical fraction (*v*) and IgG waning rate (*ω*), we refitted the model with each parameter halved and doubled in turn (*v* = 0.075 and 0.30; *ω* = 0.27 and 1.08 year^−1^), keeping all other settings unchanged (Supplementary material Text S4). Where some chains did not converge, results were summarised from the chains that mixed with each other.

### Seasonality patterns

Seasonal patterns were identified by applying *k*-means clustering to the peak months, peak multipliers and log peak-to-trough amplitude ratios derived from the posterior median seasonal multipliers (*κ*_*p,m,s*_) of each province-period. The number of clusters (*K*) was selected by comparing *K* = 1–8 using the within-cluster sum of squares (elbow method), average silhouette width^15^, and cluster stability^16^. Stability was assessed using the adjusted Rand index (ARI)^17^ to compare the clustering solution obtained from the original dataset with those obtained from 200 bootstrap resamples. To account for uncertainty in seasonal patterns, each posterior draw was assigned to the nearest cluster centroid, and the proportion of draws assigned to each cluster was used to estimate the posterior probability of pattern membership.

### Software and code availability

All analyses were conducted using R version 4.5.2. The full analysis workflow and source code for statistical modelling and visualisation are available at https://github.com/psamonwong/dynamite-506-analysis. Numerical data underlying all figures are provided in S1 Data.

## Results

### National surveillance data

Between 2003 and 2024, a total of 143,096 scrub typhus cases and 119 related deaths were reported. The five most reported provinces, most of which were located within Health Region 1, were Chiang Rai (17,135), Chiang Mai (16,324), Tak (11,864), Mae Hong Son (11,643), and Nan (9,547) (Fig 1A). Accounting for population size, these five provinces also had the highest average annual reported incidence, led by Mae Hong Son (195.0 per 100,000 population per year), followed by Tak (88.7), Nan (78.4), Chiang Rai (58.4) and Chiang Mai (40.5). At the regional level, Health Regions 1 (42%), 9 (11%), 2 (11%), 10 (11%), and 11 (7%) reported the highest cumulative case numbers, respectively. Relative to population size, reported incidence was highest in Health Regions 1 (42.4 per 100,000), 2 (18.1), 10 (13.1), 9 (9.5) and 11 (8.8), compared with 8.7 per 100,000 nationally. While cumulative deaths were not concentrated in a single health region (Fig 1B), the highest numbers were reported in Chiang Rai (29), Nakhon Ratchasima (18), Surat Thani (15), Mae Hong Son (9), and Chiang Mai (8). Relative to population size, reported mortality was highest in Mae Hong Son (1.5 per 1,000,000 population per year), Chiang Rai (1.0), Surat Thani (0.6), Phayao (0.3) and Nakhon Ratchasima (0.3). Direct age standardisation produced nearly identical rankings of provinces (Spearman correlation ≥0.98 for both incidence and mortality).

Scrub typhus cases gradually increased, reaching a peak in 2013, followed by a sharp decline until 2016 and then a subsequent rise. Extending this temporal trend, our study observed another marked decrease in reported cases during 2020–2021, likely reflecting the impact of the COVID-19 pandemic on either environmental exposure, health-seeking behaviours, or health system functioning (Fig. 1D). A similar effect was evident in mortality data, with no deaths reported in 2021 (Fig. 1E). Case reporting, however, rebounded rapidly to pre-pandemic levels from 2023 onwards. Annual reported cases and deaths in 2024 were 8,959 and 7, respectively.

Of the total reported cases, the most affected age cohorts were 45–55 (17%), 35–45 (16%), and 25–35 years (14%) (Fig 1F). In contrast, mortality was skewed towards older individuals, with the highest proportion among those aged 65 years and above (23%), followed by the 45–55 (18%) and 55–65 (16%) age groups.

### Reporting proportion

Coupling national surveillance data with seroprevalence data within a hierarchical Bayesian framework enabled estimation of reporting proportion, defined as the proportion of clinical infections reported through the surveillance system, a model-derived latent estimate rather than a directly observed surveillance metric. Reporting proportion for scrub typhus varied over time between 2003 and 2024 across all 13 Health Regions (2). During the early 2010s, Health Regions 1 and 2 showed relatively higher reporting proportions than other regions, peaking at 0.89 (95% credible interval [CrI]: 0.75–0.97) in 2013 and 0.84 (95% CrI: 0.69–0.93) in 2010, respectively. In contrast, Health Regions 4, 5, and 6 consistently exhibited lower reporting proportion, never exceeding 0.40 throughout the observation period.

A notable pattern observed across nearly all regions was a synchronized and marked decline in reporting proportion during 2020–2021 (Fig 2). In Health Region 1, reporting proportion decreased from a pre-pandemic level to 0.26 (95% CrI: 0.19 to 0.34) in 2021. Such notable reductions were observed in Health Regions 7, 9, 10, and 11, where reporting proportion fell to 0.19 (95% CrI: 0.12 to 0.29), 0.19 (95% CrI: 0.12 to 0.27), 0.20 (95% CrI: 0.12 to 0.32), and 0.33 (95%CrI: 0.24 to 0.44), respectively.

**Figure 2.**
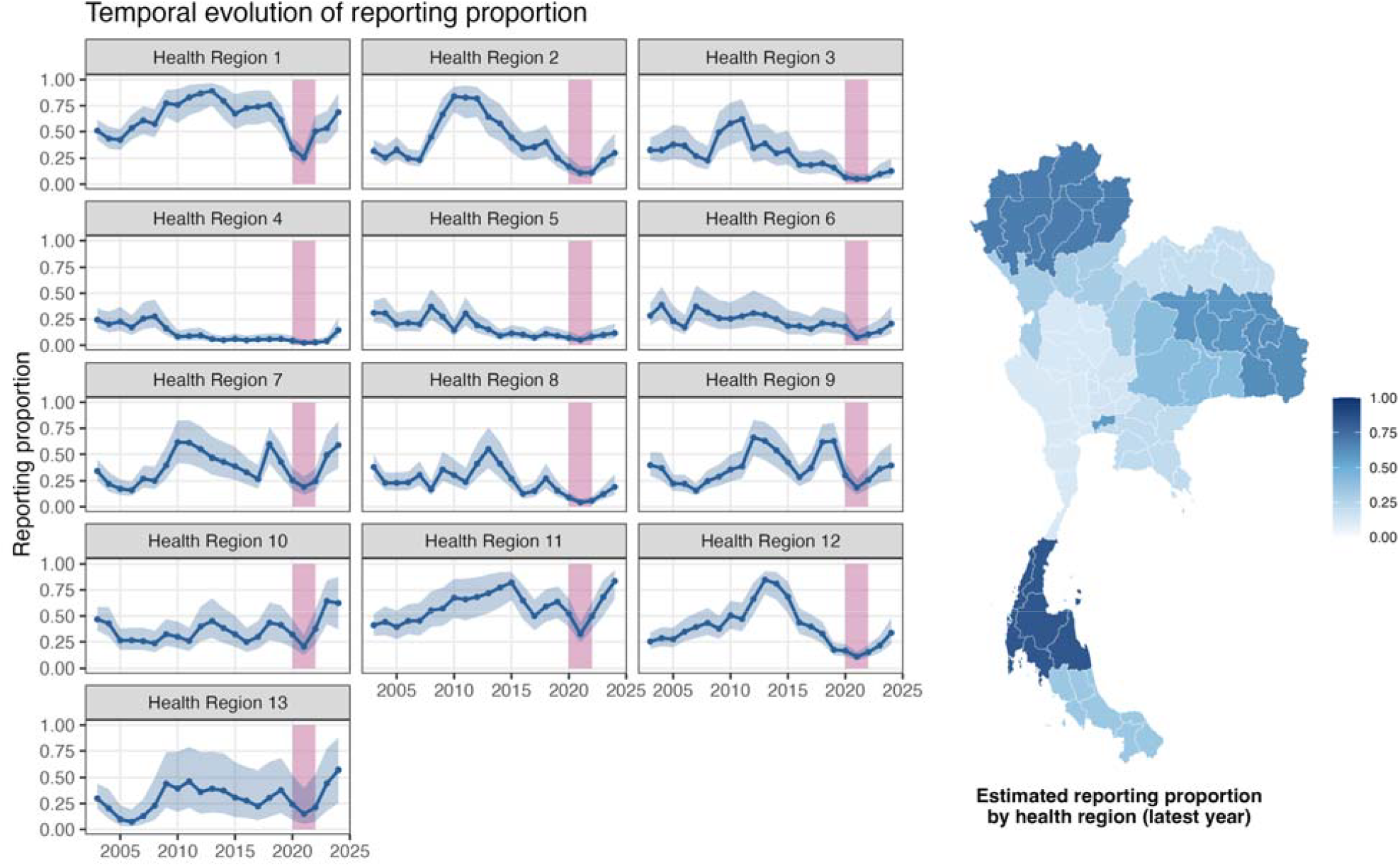
Estimated reporting proportion (π) for clinical scrub typhus by health region. Reporting proportion represents the probability that a clinical infection is diagnosed and notified as a reported case; it is a model-derived estimate rather than an observed surveillance metric. Left: Points and trend lines show median estimates, and shaded areas indicate 95% credible intervals. The pink shaded period (2020 to 2021) marks the time when the COVID-19 pandemic affected reporting proportion. Right: Map of median estimates by health region in 2024.

By 2024, all regions showed a rapid rebound in reporting proportion, albeit with some geographical variation. (Fig. 2). The spatial distribution map for 2024 indicates that the highest estimated reporting proportions were observed in Health Regions 11, 1, 10, and 7, reaching 0.83 (95% CrI: 0.66 to 0.94), 0.69 (95% CrI: 0.51 to 0.87), 0.62 (95% CrI: 0.37 to 0.87), and 0.59 (95% CrI: 0.37 to 0.82), respectively.

### Estimated force of infection and disease burden

Posterior draws showed a negative relationship between estimated force of infection and reporting proportion within province-years (Figure S1), reflecting their multiplicative contribution to reported cases. Most provinces exhibited relatively stable transmission between 2003 and 2024, with average annual FOI (the estimated annual hazard of infection per at-risk person) remaining below 10^−3^ per at-risk person-year. This corresponds to fewer than approximately 100 infections per 100,000 at-risk population per year, including both clinical and subclinical infections, particularly in Health Regions 3, 4, 5, 6, and 7 (Fig. 3). Persistent high-incidence hotspots with a gradual increasing trend were observed in northern and western border provinces, notably Mae Hong Son and Tak, where the estimated annual incidence in 2024 exceeded 1,000 infections per 100,000 at-risk population. It is worth noting that provinces in Health Regions 9 and 10 have consistently experienced high transmission throughout the observation period, with estimated FOI levels comparable to historically top-reporting provinces such as Chiang Rai, Chiang Mai, and Nan.

**Figure 3.**
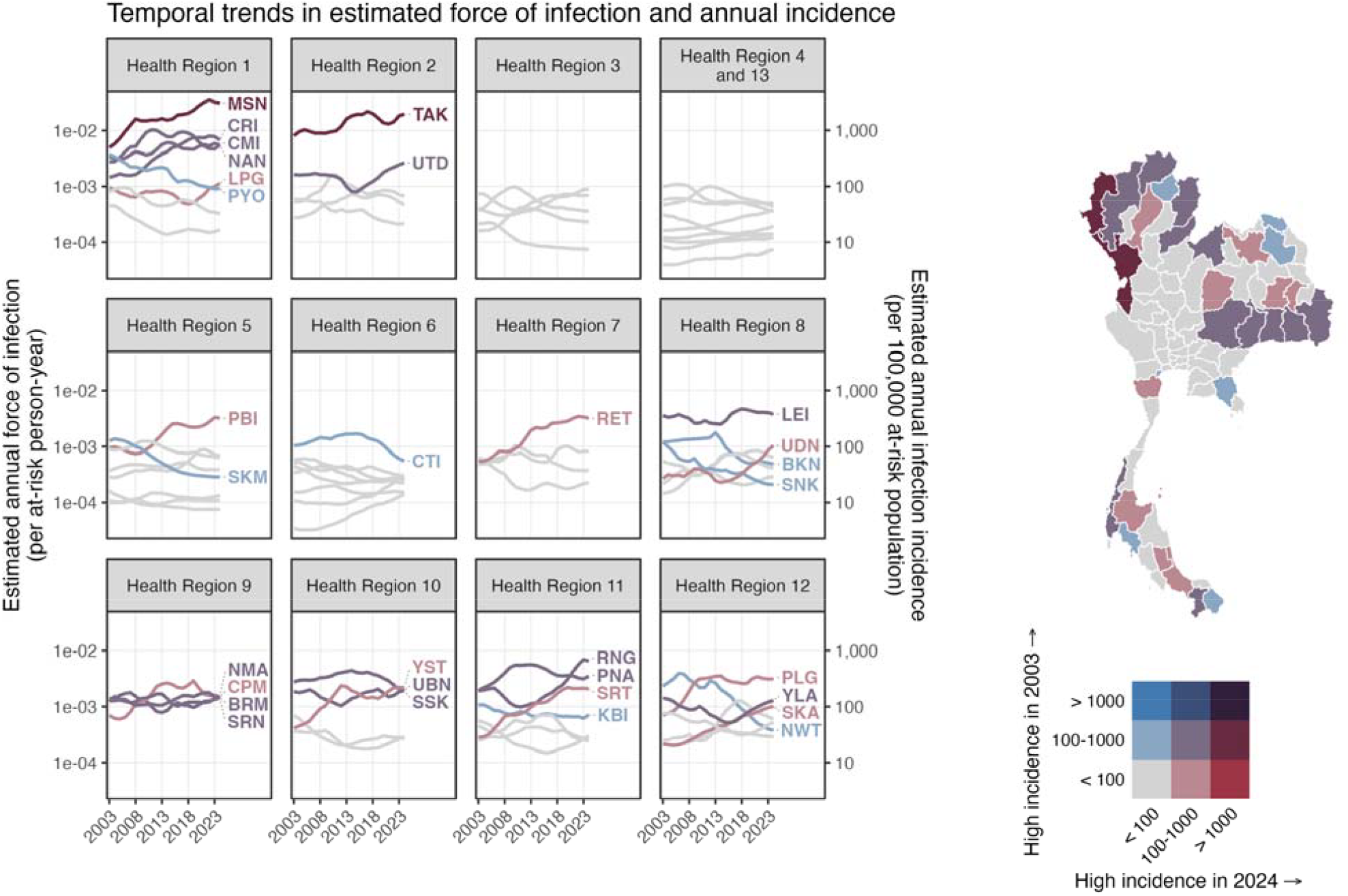
Estimated annual force of infection (FOI) and incidence by province, 2003–2024. Left: Lines represent median values for annual FOI and estimated infection incidence per 100,000 at-risk population. Colors distinguish specific temporal trend classifications. The estimates reflect all infections, including both clinical and subclinical infections, rather than reported cases. Right: Map displaying groups of provinces with contrasting incidence trends over the study period. Abbreviations: MSN, Mae Hong Son; CRI, Chiang Rai; CMI, Chiang Mai; NAN, Nan; LPG, Lampang; PYO, Phayao; TAK, Tak; UTD, Uttaradit; PBI, Phetchaburi; SKM, Samut Songkhram; CTI, Chanthaburi; RET, Roi Et; LEI, Loei; UDN, Udon Thani; BKN, Bungkan; SNK, Sakon Nakhon; NMA, Nakhon Ratchasima; SRN, Surin; CPM, Chaiyaphum; BRM, Buri Ram; YST, Yasothon; UBN, Ubon Ratchathani; SSK, Si Sa Ket; RNG, Ranong; PNA, Phang Nga; SRT, Surat Thani; KBI, Krabi; PLG, Phatthalung; SKA, Songkhla; YLA, Yala; NWT, Narathiwat.

Over the two decades, several other provinces have shown notable rising trends, including Lampang (Health Region 1), Phetchaburi (Health Region 5), Roi Et (Health Region 7), Udon Thani (Health Region 8), Chaiyaphum (Health Region 9), Yasothon (Health Region 10), Ranong, Surat Thani and Phang Nga (Health Region 11), and Phatthalung and Songkhla (Health Region 12). In contrast, declines in transmission intensity were observed in provinces such as Phayao (Health Region 1), Phra Nakhon Si Ayutthaya (Health Region 4); Samut Songkram (Health Region 5), Chanthaburi (Health Region 6), Bungkan and Sakon Nakhon (Health Region 8), Krabi (Health Region 11), and Narathiwat (Health Region 12).

### Age-specific susceptibility and case-fatality rate

Susceptibility to scrub typhus infection showed a strong association with age, particularly in the most recent periods (Fig. 4). Individuals younger than the reference age group (25–35 years) consistently exhibited lower susceptibility, with estimated relative susceptibility and 95% CrI remaining below 1. In contrast, elderly showed notable changes over time. By 2019-2024, individuals aged over 65 had a relative susceptibility of 1.6 (95% CrI: 1.5 to 1.6), compared with 0.7 (95% CrI: 0.7 to 0.8) in 2003–2007. Similar upward trends were observed in the 45–55 and 55–65 age groups across the two-decade.

**Figure 4.**
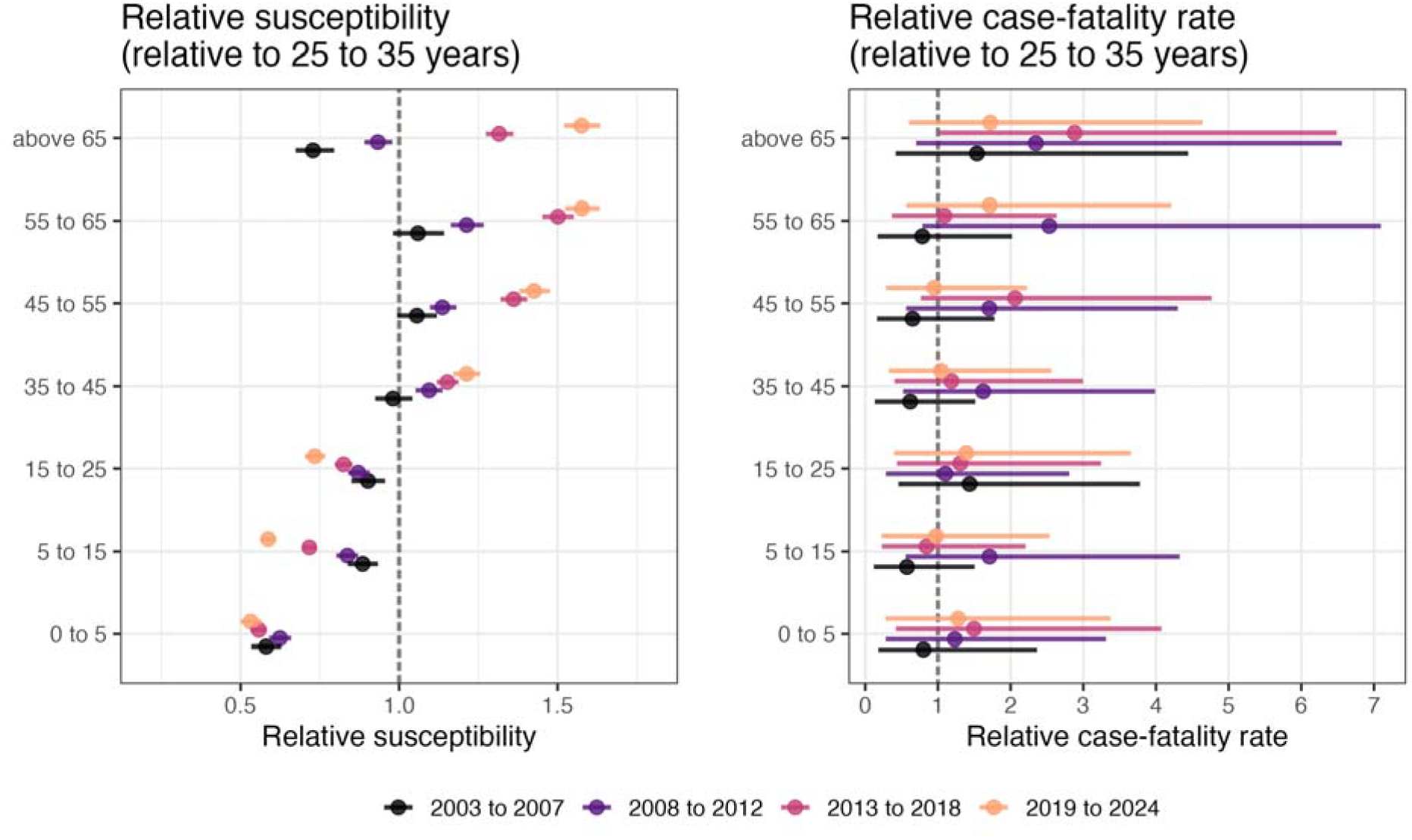
Age-specific relative infection susceptibility (*ρ)* and relative case-fatality rates among reported cases (*η*) over periods. Estimates are shown relative to the 25–35 year age group (reference). Left: Relative susceptibility to infection. Right: Relative case-fatality rate. Case-fatality is calculated among reported cases and is not an infection-fatality risk. Points and error bars represent mean estimates and 95% credible intervals, respectively.

The estimated case-fatality rate among reported cases for the reference group (25–35 years) declined sharply from 0.15% (95% CrI: 0.07–0.30%) in 2003–2007 to 0.06% (95% CrI: 0.02–0.12%) in 2008–2012, after which it remained stable at approximately 0.06– 0.07% through 2019–2024. Estimates of the relative case-fatality rate had substantially wider credible intervals than those for susceptibility, reflecting the smaller number of reported deaths (Fig. 4). Despite this uncertainty, individuals aged over 65 had consistently elevated case-fatality rates relative to the 25–35 year reference group, with posterior mean estimates exceeding 1.0 across all time periods and ranking among the highest of all age groups, although credible intervals overlapped 1 for most age groups.Combined with the patterns observed for relative susceptibility, these findings suggest that clinical infection has become increasingly concentrated among older populations over time, a group that also faces elevated mortality risk.

### Seasonal patterns

Rather than characterising seasonal patterns by administrative regions as in a previous studiy^7^, we classified seasonality at the period–province level. Using *k*-means clustering, we identified three distinct seasonal patterns, distinguished by their peak timing and peak-to-trough amplitude (Fig. 5). Three clusters corresponded to the elbow of the within-cluster sum of squares and gave the most stable partition across bootstrap resamples (median ARI 0.99 vs ≤0.84 for *K* = 4–8; Figure S2); although *K* = 2 had a higher silhouette width (0.66 vs 0.58), it was unstable (median ARI 0.29). The patterns were distinguished mainly by peak timing. Pattern 1 had a late-September peak and the highest amplitude ratio (3.1). Pattern 2 peaked in early July with a moderate amplitude ratio (2.2). Pattern 3, by contrast, exhibited relatively flat seasonality, peaking earlier in the year with the lowest amplitude ratio (1.9).

**Figure 5.**
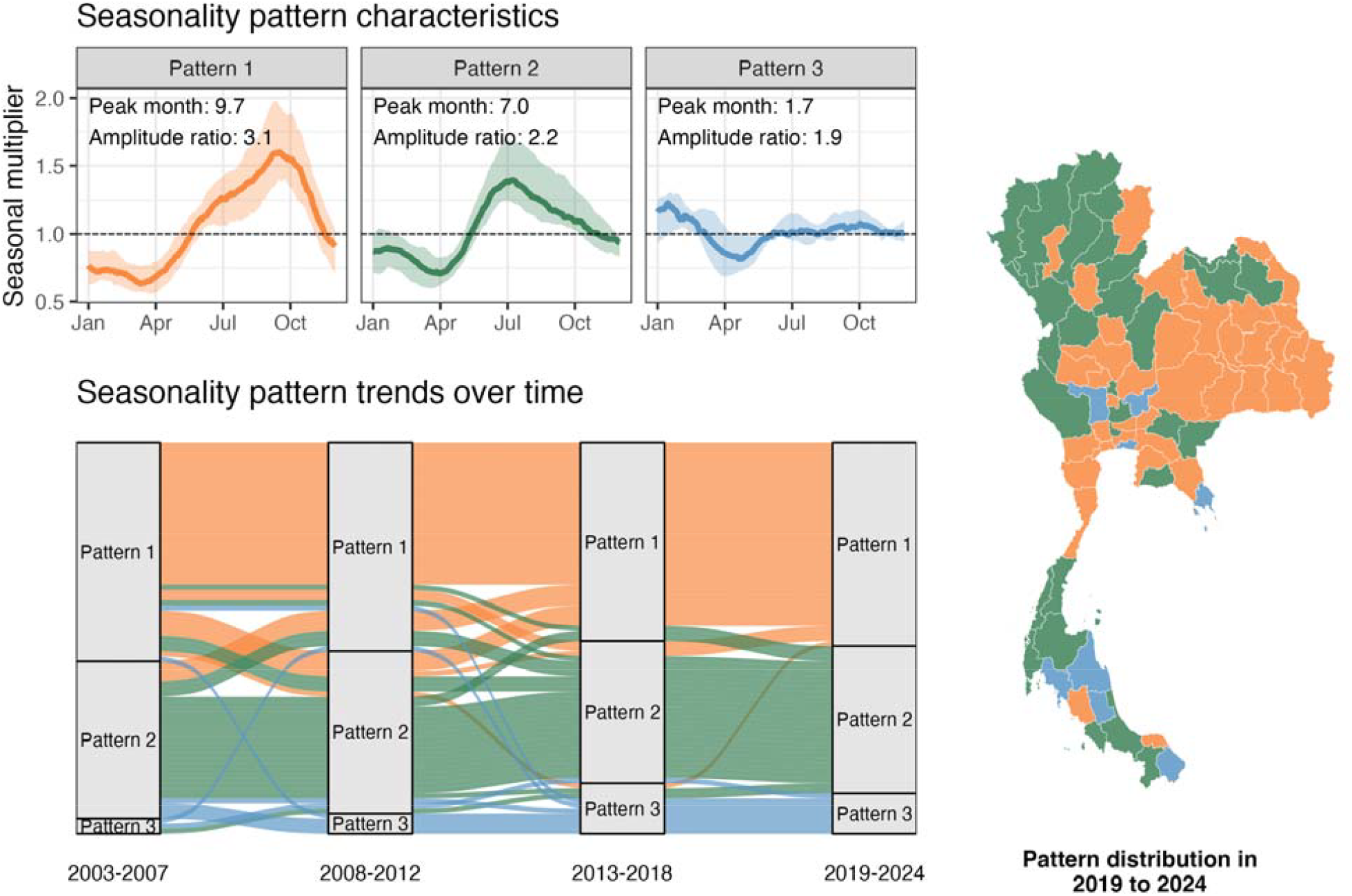
Characterization of three distinct seasonal patterns of scrub typhus, 2003–2024. Top left: Median monthly seasonal multiplier (*k*) with interquartile ranges. Bottom left: Longitudinal shifts in seasonal pattern membership across four periods; each province is colored according to its pattern assignment in the most recent period (2019–2024) to track retrospective transitions over time. Right:distribution of seasonal patterns by province for the most recent period (2019–2024).

The 2019–2024 map illustrates the geographic distribution of seasonal patterns (Fig. 5), which remained broadly stable across all periods. Pattern 1 was the most widespread, covering 52% of provinces, primarily across the Northeast and Central regions. Pattern 2 accounted for 38%, concentrated in the North, including the western border provinces, and the South. Pattern 3 represented a small but persistent share (10%), scattered across the South, East and Central regions. This stability was further reflected by the fact that 58% of provinces retained the same seasonal pattern throughout the entire observation period. When uncertainty in the estimated seasonal curves was considered, this proportion was 44% (95% CrI: 32–55%). Assignment to Pattern 3 was the least certain, reflecting its flatter seasonal profile.

## Discussion

Characterising how the epidemiology of a disease evolves over the long term is essential for assessing disease burden, guiding public health interventions, and understanding how the disease interacts with environmental and socio-economic changes, including those driven by climate. A previous study analysed nationwide passive surveillance data for scrub typhus in Thailand from 2003 to 2018, focusing on broad historical characteristics of the disease while restricting temporal trend analysis to a single province without assessing changes at the national level^7^. The present study extends these data through 2024 and examines long-term shifts over 22 years across four key aspects: (1) reporting proportions, (2) transmission intensity, (3) disease severity, and (4) seasonal patterns.

Long-term national surveillance systems offer a valuable opportunity to examine how the epidemiology of notifiable diseases evolves over space and time. In Thailand, data from the National Disease Surveillance database (R506) have been widely used to describe disease dynamics^18–21^ and, in some cases, to explore correlations with environmental variables. However, surveillance data are inherently imperfect and do not directly reflect underlying transmission. Reported incidence is shaped by the reporting proportion, which depends on diagnostic capacity, healthcare access, surveillance practices, diagnostic sensitivity and specificity, and clinical awareness. Temporal trends in these data therefore reflect a mixture of true epidemiological change and variation in detection and reporting, and may be misinterpreted if these processes are not explicitly accounted for. To address this, we integrated passive surveillance data with nationwide seroprevalence data and antibody kinetics to jointly infer reporting proportions and underlying transmission intensity.

Clinical attention to scrub typhus may have influenced case reporting. Estimated reporting proportion generally mirrored the transmission intensity across regions, with higher FOI in Health Regions 1 and 2 possibly leading clinicians to consider the disease more consistently in the differential diagnosis of undifferentiated febrile illness, thereby contributing to higher detection. By contrast, regions with lower FOI, including Health Regions 4, 5, and 6, may have given the disease less diagnostic consideration, partly explaining their lower reporting proportions due to under-diagnosis^8,22^. Beyond these regional differences, the diversion of healthcare resources during the COVID-19 pandemic (2020–2021) coincided with a temporary synchronised decline in reporting proportion. Nevertheless, the rapid rebound to near pre-pandemic levels by 2024 in most regions suggests a resilient surveillance infrastructure, likely supported by Thailand’s well-established public health system and its capacity to restore routine disease detection after major systemic disruptions. Because the reporting proportion also absorbs changes in diagnostic practice, including the gradual replacement of the Weil–Felix test by more specific assays, its temporal trends should be interpreted as the combined effect of clinical awareness, healthcare access and diagnostic performance rather than of any single factor.

After accounting for temporal variation in reporting proportions, the estimated force of infection suggested substantial geographic heterogeneity in transmission, which may reflect ecological and occupational factors such as climate, land use, and exposure to chigger habitats. Health Regions 1 and 2 in the north consistently exhibited the highest FOI, with Chiang Rai, Chiang Mai, and Nan long recognised as high-burden provinces and the setting for much of the existing field and clinical research^23–25^. Our estimates further highlight that Mae Hong Son and Tak, which have smaller populations and consequently lower absolute case counts, harbour FOI at least as high as these better-studied provinces, with both showing notable upward trends, particularly Mae Hong Son. This reinforces earlier findings by^7^, who reported that Mae Hong Son and Tak had the highest incidence rates among all Thai provinces, and suggests that these areas may benefit from increased research and public health attention. Beyond the north, the northeastern regions (Health Regions 8–10) showed moderate but sustained FOI, while the southern regions (Health Regions 11 and 12) displayed considerable within-region heterogeneity, with Ranong in particular showing a notable rise over the study period. These observations suggest that scrub typhus transmission in Thailand extends well beyond the mountainous terrain and ethnic minority populations where the disease has traditionally been focused^5,26^.

Estimated age-specific susceptibility to scrub typhus showed a temporal shift over the study period. In 2003–2007, susceptibility was relatively uniform across adult age groups, with older adults even showing lower values than the 25–35-year reference group. By 2019–2024, however, this pattern had reversed, with adults aged 55 and above showing the highest relative susceptibility. Children remained consistently less susceptible than adults throughout. This shift may reflect Thailand’s rapidly ageing agricultural workforce, in which older farmers continue to maintain exposure to chigger habitats while younger cohorts increasingly move into less exposed occupations or urban settings^27,28^. Although cumulative lifetime exposure in older adults could contribute to higher background seropositivity and potential false positive diagnoses^29^, this alone is unlikely to explain the observed temporal shift, as such an effect would be expected to remain constant across periods rather than progressively increase.

In absolute terms, case-fatality remained low throughout the study period and declined over the first decade before stabilising. This may reflect the expansion of Thailand’s Universal Coverage Scheme from 2002, which substantially improved healthcare access in rural areas where scrub typhus burden is highest^30^, enabling earlier presentation and more timely diagnosis and treatment. The relative case-fatality rate, however, was consistently elevated among adults above 65 across all periods, albeit with wide credible intervals, consistent with findings across endemic settings linking older age to higher complication rates, immunosenescence, and comorbidities^31–33^. As Thailand’s population continues to age, the combination of rising susceptibility and persistently elevated case-fatality among older adults underscores the need for targeted clinical vigilance and public health efforts in ageing rural communities.

Three seasonal patterns of scrub typhus transmission, differing mainly in peak timing, were identified across Thailand through 2003 to 2024. Pattern 1, the most common at the provincial level, showed strong seasonality peaking in late September (amplitude ratio 3.1), coinciding with the latter part of the monsoon season. Pattern 2 peaked earlier around July (amplitude ratio 2.2), while Pattern 3 showed minimal seasonal variation (amplitude ratio 1.9), suggesting near year-round transmission. Although Pattern 1 covered the majority of provinces, the national-level seasonal profile more closely resembles Pattern 2, reflecting the disproportionate contribution of high-incidence provinces that follow this earlier-peaking pattern. These patterns remained relatively stable over the two decades of the study period, with no clear directional shift between periods, suggesting that the underlying drivers of seasonality have not fundamentally changed; the shifts observed in a minority of provinces should be interpreted cautiously given uncertainty in pattern assignment, and seasonality appears to be shaped largely by relatively fixed climatic and geographic factors. In addition, while our analysis identified distinct seasonal patterns and tracked their longitudinal shifts across provinces, we did not formally investigate the role of large-scale climate variability such as the El Niño–Southern Oscillation (ENSO) in shaping these patterns. Linking annual transmission intensity at the national level to climate indices is an important next step that could help clarify the mechanisms driving observed inter-annual variability.

This subnational heterogeneity in seasonal timing is not unique to Thailand. In China, three epidemiologically distinct regions with markedly different seasonal profiles have been identified: a sharp October–November peak in the middle-eastern provinces, a broader June–September peak in the southwest, and a more extended transmission season in the southeast^34^. In South Korea, cases concentrate in October–November but the peak shifts to later months at lower latitudes^35^, while Japan shows a bimodal pattern in the north with peaks in both May and October–November, compared to a single dominant October–November peak in the south^36^. In India, peak timing varies regionally: from August to February with an October peak in the south, and broadly July to November in the north and northeast^37^. Thailand’s Pattern 3, with its near year-round transmission in the south, echoes the extended seasons seen in equatorial parts of Southeast China and Yunnan, where year-round rainfall likely sustains more constant mite populations.

Taken together, these comparisons suggest that scrub typhus seasonality is shaped by the interplay of mite species composition, host population dynamics, human exposure patterns linked to agricultural calendars and land use, and abiotic conditions that influence all of these simultaneously. Predicting case reports directly from climate data alone, as several studies have attempted, has therefore limitations, not only because climate acts indirectly through these intermediary pathways, but also because reported case counts are further filtered through healthcare-seeking behaviour, diagnostic practices, and reporting systems that may themselves vary seasonally.

Given this complexity, effective control measures, whether through targeted vector management, seasonally timed risk communication and prevention, or clinical preparedness, need to be locally tailored to the specific drivers of transmission in each setting. This requires disentangling the contributions of each of these interacting factors, yet several critical data gaps remain to be addressed.

Despite Thailand being a major endemic setting, its chigger vector ecology remains largely unexplored. More than 20 *Leptotrombidium* species have been reported from Thailand^38^, and *O. tsutsugamushi* has been recently detected in chiggers from multiple genera beyond *Leptotrombidium*^23^, yet vector competence, geographic distribution, and seasonal activity have been studied for only a handful of species, mostly in the northern provinces and only for short periods. Whether the distinct seasonal patterns identified in our study correspond to different dominant vector species, as demonstrated in Japan, South Korea, and China^35,36,39^, or instead reflect regional differences in agricultural calendars and human exposure, remains an open question.

Beyond vector ecology, important gaps remain in our understanding of rodent host dynamics and their overlap with infected vectors, and occupational and behavioural exposure patterns across regions. Mathematical modelling frameworks that integrate vector, host, and human data could help translate these observations into locally appropriate strategies. Realising this will require sustained and standardised surveillance across all three dimensions, both to inform intervention design and to evaluate effectiveness over time.

As with any modelling study that relies on passively collected surveillance data, our approach is subject to certain limitations that should be considered when interpreting the findings. First, baseline force of infection and reporting proportions were estimated jointly and cannot be fully separated using surveillance data alone, since reported cases depend on both. The model better identifies relative spatial differences because reporting proportion varies by region, whereas transmission varies by province. Seroprevalence data anchor the overall level only in 2007, 2008 and 2012; otherwise, separation relies on gradually changing reporting. We therefore interpret spatial patterns and relative differences more confidently than absolute estimates. The 2020–2021 decline in reported cases is robust, but its attribution to reduced detection versus reduced transmission remains uncertain. Second, some model parameters, such as antibody kinetics and the clinical fraction, were informed by published literature rather than estimated directly from the data, and may not fully reflect the local epidemiological context in Thailand. We assessed the influence of these assumptions by refitting the model with each parameter halved and doubled (Supplementary material Text S4;Figures S3–S5). Doubling the clinical fraction halved the estimated force of infection but left the ranking of provinces, reporting proportions, age-specific susceptibility and case-fatality estimates essentially unchanged, and halving the waning rate had a negligible effect. With a halved clinical fraction, two of four chains did not converge and were excluded, and the remaining chains showed the same pattern with an approximately doubled force of infection. With a doubled waning rate, no two chains converged, and the most consistent chains implied implausibly intense transmission and very low reporting proportions, suggesting that such rapid antibody waning is difficult to reconcile with the observed data. Because these parameters scale the relationship between infections and reported cases, they affect absolute estimates more than relative comparisons between provinces or age groups. Third, the model assumed uniform antibody dynamics across age groups, without accounting for potential age-specific differences in serological response, such as slower antibody decay or higher baseline titres in older adults with repeated lifetime exposure. Incorporating age-specific serodynamics could refine estimates of susceptibility and help distinguish genuine changes in exposure from diagnostic artefacts. A further consideration is that the seroprevalence data used for calibration were collected from young men, whose exposure patterns are unlikely to be representative of children, women or older adults; age-specific estimates outside this range therefore depend more heavily on the model structure than on direct serological observation. Although the serological data were linked to the 15–25-year age group, the model does not distinguish between sexes; if young men are more exposed than young women, as is plausible given occupational and outdoor activities, the estimated force of infection may be overestimated and reporting proportions correspondingly underestimated. Fourth, the model did not explicitly estimate false positive rates; in endemic settings, background seropositivity from past infections can lead to misclassification, particularly among older adults with cumulative lifetime exposure, potentially inflating apparent susceptibility in these age groups. Fifth, age-specific susceptibility and case-fatality were estimated at the national level and did not explore regional variation, which may be substantial given the geographic heterogeneity in transmission intensity, bacterial strains, occupational structure, and healthcare access observed across Thailand. Sixth, surveillance case definitions combined suspected, probable and confirmed cases, and diagnostic practice shifted over the study period from serological methods with limited specificity towards more specific assays. The aggregated data did not record case classification or the diagnostic test used, precluding their inclusion as covariates in the reporting model. Changes in diagnostic practice may therefore contribute to the estimated temporal trends in reporting, which we could not separate from changes in healthcare-seeking or surveillance capacity. However, because the reporting proportion is shared by provinces within each health region, region-wide changes in diagnostic practice cannot explain the contrasting transmission trends observed among provinces within the same region, such as Phatthalung and Narathiwat in Health Region 12, although differences in diagnostic practice between provinces could. Finally, case-fatality estimates are conditional on reported cases and therefore reflect outcomes among individuals who sought healthcare and received a diagnosis, rather than among all infected individuals; if healthcare-seeking behaviour or diagnostic sensitivity varies by age or region, the estimated rates may not be representative of the true infection fatality.

## Conclusion

In conclusion, our model-based estimates suggest that scrub typhus transmission in Thailand is not confined to the north, and that the northeastern and southern regions may represent substantial and under-recognised foci, with estimated force of infection rising in several provinces. While passive surveillance data are inherently shaped by reporting practices that vary across regions and time, Thailand’s long-standing national surveillance infrastructure provides a valuable resource for tracking the evolving epidemiology of scrub typhus when these biases are explicitly modelled and estimates are interpreted in light of their assumptions. The disease burden is increasingly concentrated among older adults, who show both rising susceptibility and elevated case-fatality. Seasonal timing of transmission varies markedly across regions but has remained largely stable over two decades. Together, these findings highlight the need to broaden surveillance and research to underserved endemic areas, direct clinical awareness and public health efforts toward ageing rural populations, and design interventions informed by the distinct seasonal and ecological conditions of each region. This will require deeper understanding of the interacting vector, host, and human drivers that shape transmission locally.

## Data Availability

The data underlying this article are available in Github at https://github.com/psamonwong/dynamite-506-analysis.

## Supplementary material

Supplementary manterial is available at IJE online.

## Acknowledgments

The authors would like to express their gratitude to the Department of Disease Control, Ministry of Public Health of Thailand for kindly providing the data used in this study.

## Author contributions

Conceptualization: Phrutsamon Wongnak. Data curation: Phrutsamon Wongnak. Formal analysis: Phrutsamon Wongnak. Funding acquisition: Wirichada Pan-ngum. Investigation: Phrutsamon Wongnak. Methodology: Phrutsamon Wongnak. Project administration: Wirichada Pan-ngum. Resources: Darin Areechokchai. Software: Phrutsamon Wongnak. Supervision: Wirichada Pan-ngum. Visualization: Phrutsamon Wongnak. Writing – original draft: Phrutsamon Wongnak. Writing – review & editing: Phrutsamon Wongnak, Kittipong Chaisiri, Carlo Perrone, Karine Chalvet-Monfray, Darin Areechokchai, Wirichada Pan-ngum. All authors approved the final version of the manuscript.

## Data availability

All data and code underlying this article are publicly available. The processed input data and the full analysis code are available at https://github.com/psamonwong/dynamite-506-analysis. The numerical data underlying all figures, including province-level annual reported cases, deaths and incidence, estimated force of infection and infection incidence, reporting proportions by health region and year, age-specific susceptibility and case-fatality estimates, seasonal patterns, cluster validity metrics and the results of the sensitivity analysis, are provided in S1 Data and in the Source data folder of the repository. Posterior predictive checks and MCMC trace plots are available at https://doi.org/10.5281/zenodo.23941245.

## Use of Artificial Intelligence (AI) Tools

OpenAI’s GPT-5.3 model was used exclusively for language editing to improve the grammar and clarity of the manuscript and for commenting and annotating programming code. The authors were solely responsible for all scientific aspects of this work, including conceptualisation, study design, data analysis, interpretation of results, and figure generation. The authors reviewed and take full responsibility for all final content.

## Supporting information captions

**S1 Text**. Supplementary model descriptions.

**S2 Text**. Prior specifications.

**S3 Files**. Posterior predictive checks and traceplots.

**S4 Text**. Sensitivity analysis for the clinical fraction and IgG waning rate.

**S1 Data**. Numerical data underlying Figs 1–5, Figures S2–S5 and Table S4.1.

## References

1. Trishnananda M, Vasuvat C, Harinasuta C. Investigation of scrub typhus in thailand. Journal of Tropical Medicine and Hygiene. London; 1964;67(9):215–219 pp.

2. Wangrangsimakul T, Greer RC, Chanta C, et al. Clinical characteristics and outcome of children hospitalized with scrub typhus in an area of endemicity. Journal of the Pediatric Infectious Diseases Society. 2020 Apr;9(2):202–209.

3. Singh P. Scrub typhus, a case report : Military and regional significance. Medical Journal Armed Forces India. 2004 Jan;60(1):89–90.

4. Mathai E, Rolain JM, Verghese GM, et al. Outbreak of scrub typhus in southern india during the cooler months. Ann N Y Acad Sci. United States; 2003 June;990:359–364.

5. Tasak N, Apidechkul T, Law ACK, et al. Prevalence of and factors associated with scrub typhus exposure among the hill tribe population living in high incidence areas in thailand: A cross-sectional study. BMC Public Health. England; 2023 Dec;23(1):2394.

6. Elliott I, Pearson I, Dahal P, Thomas NV, Roberts T, Newton PN. Scrub typhus ecology: A systematic review of orientia in vectors and hosts. Parasites & Vectors. 2019 Nov;12(1):513.

7. Wangrangsimakul T, Elliott I, Nedsuwan S, et al. The estimated burden of scrub typhus in thailand from national surveillance data (2003-2018). PLOS Neglected Tropical Diseases. Public Library of Science; 2020 Apr;14(4):e0008233.

8. Gibbons CL, Mangen M-JJ, Plass D, et al. Measuring underreporting and under-ascertainment in infectious disease datasets: A comparison of methods. BMC Public Health. England; 2014 Feb;14:147.

9. Division of Epidemiology, Department of Disease Control. Scrub typhus [Internet]. 2026. Available from: http://doe1.moph.go.th/surdata/disease.php?dcontent=old&ds=44

10. Division of Epidemiology, Department of Disease Control. Case definition for communicable diseases surveillance, thailand, 2020 [Internet]. Nonthaburi: Division of Epidemiology, Department of Control (TH); 2020 [cited 2025 Nov 28]. Available from: https://ddc.moph.go.th/uploads/publish/1142920210518092542.pdf

11. Gonwong S, Mason CJ, Chuenchitra T, et al. Nationwide seroprevalence of scrub typhus, typhus, and spotted fever in young thai men. Am J Trop Med Hyg. United States; 2022 Apr;106(5):1363–1369.

12. Bureau of Registration Administration. Administrative information system: Population statistics by province [Internet]. 2026. Available from: https://stat.bora.dopa.go.th/StatMIS/#/ReportStat/3

13. Devamani C, Alexander N, Chandramohan D, et al. Incidence of scrub typhus in rural south india. New England Journal of Medicine [Internet]. 2025;392(11):1089–1099. Available from: https://www.nejm.org/doi/full/10.1056/NEJMoa2408645

14. Aiemjoy K, Katuwal N, Vaidya K, et al. Estimating the seroincidence of scrub typhus using antibody dynamics after infection. Am J Trop Med Hyg. United States; 2024 June;111(2):267–276.

15. Rousseeuw PJ. Silhouettes: a graphical aid to the interpretation and validation of cluster analysis. Journal of Computational and Applied Mathematics. 1987;20:53–65.

16. Hennig C. Cluster-wise assessment of cluster stability. Computational Statistics & Data Analysis. 2007;52(1):258–271.

17. Hubert L, Arabie P. Comparing partitions. Journal of Classification. 1985;2(1):193–218.

18. Sawangpol C, Aimyong N, Phosri A. Epidemiological changes in the incidence of human leptospirosis in thailand: Findings from the national disease surveillance system from 2013 to 2022. Infectious Diseases Now. 2025 Sept;55(6):105108.

19. Chaidilok P, Saita S. Spatial and temporal analysis of dengue incidence in northern thailand: A 13-year retrospective study (2012–2024). Discover Public Health. 2026 Feb;23(1):206.

20. Kerdsin A. Human streptococcus suis infections in thailand: Epidemiology, clinical features, genotypes, and susceptibility. Tropical Medicine and Infectious Disease. 2022;7(11):359.

21. Abdulsalam FI, Antúnez P, Jawjit W. Spatio-temporal dengue risk modelling in the south of thailand: A bayesian approach to dengue vulnerability. PeerJ. United States; 2023 July;11:e15619.

22. Durrheim DN, Thomas J. General practice awareness of notifiable infectious diseases. Public Health [Internet]. 1994;108(4):273–278. Available from: https://www.sciencedirect.com/science/article/pii/S0033350694800065

23. Elliott I, Thangnimitchok N, Chaisiri K, et al. Orientia tsutsugamushi dynamics in vectors and hosts: Ecology and risk factors for foci of scrub typhus transmission in northern thailand. Parasites & Vectors. 2021 Oct;14(1):540.

24. Linsuwanon P, Auysawasdi N, Wongwairot S, et al. Assessing scrub typhus and rickettsioses transmission risks in the chiang rai province of northern thailand. Travel Medicine and Infectious Disease. 2021 July;42:102086.

25. Wangrangsimakul T, Althaus T, Mukaka M, et al. Causes of acute undifferentiated fever and the utility of biomarkers in chiangrai, northern thailand. PLOS Neglected Tropical Diseases. Public Library of Science; 2018 May;12(5):e0006477.

26. Perrone C, Kanthawang N, Cheah PY, et al. Community engagement around scrub typhus in northern thailand: A pilot project. Trans R Soc Trop Med Hyg. England; 2024 Oct;118(10):666–673.

27. Jansuwan P, Zander KK. What to do with the farmland? Coping with ageing in rural thailand. Journal of Rural Studies. 2021 Jan;81:37–46.

28. Szabo S, Apipoonanon C, Pramanik M, Leeson K, Singh DR. Perceptions of an ageing agricultural workforce and farmers’ productivity strategies: Evidence from prachinburi province, thailand. Outlook Agric. SAGE Publications Ltd; 2021 Sept;50(3):294–304.

29. Varghese GM, Rajagopal VM, Trowbridge P, Purushothaman D, Martin SJ. Kinetics of IgM and IgG antibodies after scrub typhus infection and the clinical implications. International Journal of Infectious Diseases. 2018 June;71:53–55.

30. Tangcharoensathien V, Witthayapipopsakul W, Panichkriangkrai W, Patcharanarumol W, Mills A. Health systems development in thailand: A solid platform for successful implementation of universal health coverage. The Lancet. Elsevier; 2018 Mar;391(10126):1205–1223.

31. Jang M-O, Kim JE, Kim UJ, et al. Differences in the clinical presentation and the frequency of complications between elderly and non-elderly scrub typhus patients. Archives of Gerontology and Geriatrics. 2014 Mar;58(2):196–200.

32. Taylor AJ, Paris DH, Newton PN. A systematic review of mortality from untreated scrub typhus (orientia tsutsugamushi). PLOS Neglected Tropical Diseases. Public Library of Science; 2015 Aug;9(8):e0003971.

33. Peng P-Y, Xu L, Sun J-Q, et al. Epidemiologic features and potential year of life lost of scrub typhus in china: A nationwide surveillance analysis (2006–2023). PLOS Neglected Tropical Diseases. Public Library of Science; 2025 Oct;19(10):e0013666.

34. Li Z, Xin H, Sun J, et al. Epidemiologic changes of scrub typhus in china, 1952-2016. Emerg Infect Dis. United States; 2020 June;26(6):1091–1101.

35. Jeung YS, Kim C-M, Yun NR, Kim S-W, Han MA, Kim D-M. Effect of latitude and seasonal variation on scrub typhus, south korea, 2001-2013. Am J Trop Med Hyg. United States; 2015 Oct;94(1):22–25.

36. Kinoshita H, Arima Y, Shigematsu M, et al. Descriptive epidemiology of rickettsial infections in japan: Scrub typhus and japanese spotted fever, 2007-2016. Int J Infect Dis. Canada; 2021 Feb;105:560–566.

37. D’Cruz S, Sreedevi K, Lynette C, Gunasekaran K, Prakash JAJ. Climate influences scrub typhus occurrence in vellore, tamil nadu, india: Analysis of a 15-year dataset. Scientific Reports. 2024 Jan;14(1):1532.

38. Chaisiri K, Stekolnikov AA, Makepeace BL, Morand S. A revised checklist of chigger mites (acari: Trombiculidae) from thailand, with the description of three new species. Journal of Medical Entomology. 2016 Mar;53(2):321–342.

39. Lv Y, Guo X, Jin D, et al. Infestation and seasonal fluctuation of chigger mites on the southeast asian house rat (rattus brunneusculus) in southern yunnan province, china. International Journal for Parasitology: Parasites and Wildlife. 2021 Apr;14:141–149.

