## Supplementary for "The evolving epidemiology of scrub typhus in Thailand (2003–2024): insights from latent process modelling of national surveillance data"

**This file includes:**

- Supplementary information Text S1: Supplementary model descriptions
- Supplementary information Text S2: Prior specifications
- Supplementary information files: Posterior predictive checks and traceplots
- Supplementary information Text S4: Sensitivity analysis for the clinical fraction and IgG waning rate
- Supplementary figures S1–S5

### Supplementary information Text S1: Supplementary model descriptions

#### S1.1 Random walk process for reporting proportion $\boldsymbol{\pi}_{\boldsymbol{y}\mathbf{,}\boldsymbol{r}}$

The reporting proportion $\pi_{y,r}$ was parameterized on the logit scale, ensuring values remain bounded in $[0,1]$, and was allowed to evolve over years as a region-specific Gaussian random walk:

$$\begin{matrix} \begin{aligned} \text{logit}(\pi_{1,r})&=\alpha_{r}^{\pi}, \\ \text{logit}(\pi_{y,r})&=\text{logit}(\pi_{y-1,r})+\epsilon_{y,r}^{\pi}, y=2,\ldots,Y, \\ \epsilon_{y,r}^{\pi}&\mathcal{\sim N(}0,\sigma_{\pi}^{2}), \end{aligned} \end{matrix}$$

(S1)

where $\alpha_{r}^{\pi}$ is a region-specific initial value, $\epsilon_{y,r}^{\pi}$ is the annual innovation representing the year-to-year change in reporting for region $r$, and $\sigma_{\pi}$ is the standard deviation governing the magnitude of these annual changes.

#### S1.2 Random walk process for baseline force of infection $\boldsymbol{\lambda}_{\boldsymbol{y}\mathbf{,}\boldsymbol{s}}^{\text{base}}$

The baseline force of infection $\lambda_{y,s}^{\text{base}}$ represents the average monthly transmission intensity in province $s$ during year $y$. It was parameterized on the log scale as:

$$\begin{matrix} \log\lambda_{y,s}^{\text{base}}=log\lambda_{0,s}+\epsilon_{y,s}^{\lambda}, \end{matrix}$$

(S2)

where $\log\lambda_{0,s}\mathcal{\sim N(}\mu_{\log\lambda_{0}},\sigma_{\lambda_{0}}^{2})$ is a province-specific intercept centered on a single global mean $\mu_{\log\lambda_{0}}$, and $\epsilon_{y,s}^{\lambda}$ is a cumulative random walk:

$$\begin{aligned} \epsilon_{1,s}^{\lambda}&\mathcal{\sim N(}0,\sigma_{\lambda,\text{init}}^{2}), \\ \epsilon_{y,s}^{\lambda}&=\epsilon_{y-1,s}^{\lambda}+\eta_{y,s}^{\lambda}, \eta_{y,s}^{\lambda}\mathcal{\sim N(}0,\sigma_{\lambda,y}^{2}). \end{aligned}$$

(S3)

The intercept sets the long-term transmission level for each province, while the random walk captures gradual drift over time.

#### S1.3 Fourier series for seasonality modifier $\boldsymbol{\kappa}_{\boldsymbol{p}\mathbf{,}\boldsymbol{m}\mathbf{,}\boldsymbol{r}}$

Seasonal variation was modeled using a Fourier series with $H=3$ harmonics. The log seasonal multiplier for month $m$, period $p$, and province $s$ is:

$$\begin{matrix} \log\kappa_{p,m,s}=\sum_{h=1}^{H} \left[ a_{h,p,s}^{\cos}\cos\left( {2\pi hm}/{12} \right)+a_{h,p,s}^{\sin}\sin\left( {2\pi hm}/{12} \right) \right]. \end{matrix}$$

(S4)

The coefficients were constructed using a non-centered parameterization that separates a province-level baseline (shared across periods) from period-specific deviations:

$$\begin{matrix} a_{h,p,s}^{\cos}=\tau_{s}^{\cos}\cdot\tilde{a}_{h,s}^{\cos}+\sigma_{\delta,p}^{\cos}\cdot\delta_{h,p,s}^{\cos}, \end{matrix}$$

(S5)

and analogously for the sine terms. Here $\tilde{a}_{h,s}^{\cos}$ is the province baseline coefficient, $\tau_{s}^{\cos}$ controls province-level amplitude, and $\delta_{h,p,s}^{\cos}$ is a standardized period deviation scaled by $\sigma_{\delta,p}^{\cos}$. The period SDs evolved as a random walk across periods to enforce gradual change in seasonal shape. Three harmonics were selected based on posterior predictive checks.

### Supplementary information Text S2: Prior specifications

All parameters were assigned weakly informative priors to provide broad regularization while allowing the data to dominate posterior inference. Table S2.1 summarises the prior distributions for all estimated parameters, and Table S2.2 lists the fixed hyperparameters and externally sourced values. Variance components for the force of infection random walk ($\sigma_{\lambda_{0}}$, $\sigma_{\lambda,\text{init}}$, $\sigma_{\lambda,y}$) were fixed rather than estimated following exploratory fits that revealed weak joint identifiability when all three were estimated simultaneously. Tight priors on $\sigma_{\pi}$ and $\sigma_{\rho}$ were similarly adopted to prevent degenerate behavior identified during model development.

Table S2.1 Prior distributions for estimated model parameters. $\mathcal{N}_{\boldsymbol{+}}$ denotes a normal distribution truncated to the positive real line.

| **Parameter** | **Symbol** | **Prior** |
| --- | --- | --- |
| **Observation model** | | |
| Case dispersion | $\phi_{c}$ | $\mathcal{N(}5, 2^{2}), \phi_{c}>0$ |
| Death dispersion | $\phi_{d}$ | $\mathcal{N(}3, 2^{2}), \phi_{d}>0$ |
| **Reporting proportion** | | |
| Initial value | $\alpha_{r}^{\pi}$ | $\mathcal{N(}\text{logit}(0.4), {0.3}^{2})$ |
| Annual innovation | $\epsilon_{y,r}^{\pi}$ | $\mathcal{N(}0, \sigma_{\pi}^{2})$ |
| Innovation SD | $\sigma_{\pi}$ | $\mathcal{N}_{+}(0.6, {0.01}^{2})$ |
| **Force of infection** | | |
| Global mean | $\mu_{\log\lambda_{0}}$ | $\mathcal{N(-}10, {0.5}^{2})$ |
| Province intercept | $\log\lambda_{0,s}$ | $\mathcal{N(}\mu_{\log\lambda_{0}}, \sigma_{\lambda_{0}}^{2})$ |
| Initial innovation | $\epsilon_{s}^{\lambda,\text{init}}$ | $\mathcal{N(}0, \sigma_{\lambda,\text{init}}^{2})$ |
| Annual innovation | $\eta_{y,s}^{\lambda}$ | $\mathcal{N(}0, \sigma_{\lambda,y}^{2})$ |
| **Seasonality** | | |
| Baseline Fourier coefficients | $\tilde{a}_{h,s}^{\cos}, \tilde{a}_{h,s}^{\sin}$ | $\mathcal{N(}0, 1)$ |
| Province amplitude scales | $\tau_{s}^{\cos}, \tau_{s}^{\sin}$ | $\mathcal{N}_{+}(0, {0.5}^{2})$ |
| Period deviations (raw) | $\delta_{h,p,s}^{\cos}, \delta_{h,p,s}^{\sin}$ | $\mathcal{N(}0, 1)$ |
| Initial period SD | $\sigma_{\delta,1}^{\cos}, \sigma_{\delta,1}^{\sin}$ | $\mathcal{N}_{+}(0, {0.05}^{2})$ |
| Subsequent period SD | $\sigma_{\delta,p}^{\cos}, \sigma_{\delta,p}^{\sin}$ | $\mathcal{N}_{+}(\sigma_{\delta,p-1}^{\cdot}, {0.05}^{2})$ |
| **Age-specific susceptibility** | | |
| Log-susceptibility | $\log\rho_{p,a}$ | $\mathcal{N(}0, \sigma_{\rho}^{2})$ |
| Susceptibility SD | $\sigma_{\rho}$ | $\mathcal{N}_{+}(0.3, {0.01}^{2})$ |
| **Case-fatality rate** | | |
| Mean logit-CFR | $\mu_{\text{logit-CFR}}$ | $\mathcal{N(}\text{logit}(0.05), 1^{2})$ |
| Raw deviations | $z_{p,a}^{\text{CFR}}$ | $\mathcal{N(}0, 1)$ |
| CFR SD | $\sigma_{\text{CFR}}$ | $\text{Exp}(1)$ |
| **Seroprevalence** | | |
| Pre-2003 cumulative hazard | $H_{0,s}$ | $\text{Exp}(1)$ |

Table S2.2 Fixed hyperparameters and external parameters.

| **Parameter** | **Symbol** | **Value** | **Source** |
| --- | --- | --- | --- |
| Clinical fraction | $\nu$ | 0.15 | [1] |
| IgG waning rate | $\omega$ | 0.54 yr${}^{-1}$ | [2] |
| Seroprevalence SD | $\sigma_{\text{sero}}$ | 0.01 | Calibrated |
| FOI between-province SD | $\sigma_{\lambda_{0}}$ | 0.1 | Calibrated |
| FOI initial innovation SD | $\sigma_{\lambda,\text{init}}$ | 0.75 | Calibrated |
| FOI annual innovation SD | $\sigma_{\lambda,y}$ | 0.122 | Calibrated |
| Fourier harmonics | $H$ | 3 | Posterior predictive checks |
| Serology age group | $a^{\mathrm{sero}}$ | 15–24 yr | Age of surveyed population [3] |
| Reference age group | $a^{*}$ | 25–34 yr | By design |

### Supplementary information S3: Posterior predictive checks and traceplots

Province-specific posterior predictive checks and Monte Carlo Markov Chain traceplots can be found at <https://doi.org/10.5281/zenodo.23941245>.

### Supplementary information Text S4: Sensitivity analysis for the clinical fraction and IgG waning rate

The clinical fraction ($\nu$ = 0.15) and the IgG waning rate ($\omega$ = 0.54 $\mathrm{year}^{-1}$) were fixed at values from the literature (Table S2.2). To assess their influence, we refitted the model with each parameter halved and doubled in turn while holding the other at its baseline value ($\nu$ = 0.075 and 0.30; $\omega$ = 0.27 and 1.08 $\mathrm{year}^{-1}$). All other settings, including data, priors, sampler settings and random seed, were unchanged. Convergence was assessed with the rank-normalised $\hat{R}$ statistic for key parameters. Where some chains did not converge, results were summarised from the largest set of chains that mixed with each other (maximum $\hat{R}$ < 1.1); where no two chains mixed, the two most consistent chains were used and the scenario is reported as not converged.

The refits with $\nu$ = 0.30 and $\omega$ = 0.27 $\mathrm{year}^{-1}$ converged in all four chains (maximum $\hat{R}$ 1.05 and 1.07). Doubling the clinical fraction halved the estimated force of infection (geometric mean ratio 0.50 relative to baseline), because fewer infections are needed to generate the same number of clinical cases, but the ranking of provinces by force of infection (Spearman correlation 1.00), reporting proportions (ratio 1.00), age-specific relative susceptibility and case-fatality estimates were essentially unchanged (Table S4.1; Figures S3–S5). Halving the waning rate had a negligible effect on all estimates.

For $\nu$ = 0.075, two of the four chains did not converge and were excluded; the remaining two chains mixed well (maximum $\hat{R}$ 1.02). Halving the clinical fraction approximately doubled the estimated force of infection (ratio 2.01), while the ranking of provinces (Spearman correlation 1.00), reporting proportions (ratio 0.99), the increase in relative susceptibility among adults aged over 65 (from 0.73 in 2003–2007 to 1.58 in 2019–2024, as at baseline) and case-fatality estimates were unchanged.

For $\omega$ = 1.08 $\mathrm{year}^{-1}$, corresponding to IgG remaining above the seropositivity cut-off for about 11 months on average, no two chains converged (maximum $\hat{R}$ 1.99 across all chains). Each chain was individually stationary, but the chains differed in the force of infection estimated for some provinces. Results from the two most consistent chains (maximum $\hat{R}$ 1.37) are reported for completeness but should not be interpreted quantitatively. In this scenario, the observed seroprevalence could be reproduced only with a force of infection around 200 times higher than at baseline and reporting proportions below 1%, implying implausibly intense transmission given the number of reported cases; this scenario also exaggerated the shift in susceptibility towards older adults.

Overall, the clinical fraction scales the absolute force of infection and incidence approximately in inverse proportion, whereas relative comparisons between provinces and age groups, reporting proportions and case-fatality estimates were robust across the converged scenarios. Absolute estimates of transmission should therefore be interpreted in light of these assumptions.

Table S4.1 Convergence and key estimates under each sensitivity scenario.

| **Scenario** | **Chains used** | **Max** $\hat{R}$ | **FOI ratio** | **Spearman (FOI)** | **Reporting ratio** | $\rho$ **>65, 2003–2007** | $\rho$ **>65, 2019–2024** | **CFR 25–35 y, 2003–2007 (%)** |
| --- | --- | --- | --- | --- | --- | --- | --- | --- |
| Baseline ($\nu$ = 0.15, $\omega$ = 0.54) | 4/4 | 1.02 | 1.00 | 1.00 | 1.00 | 0.73 | 1.58 | 0.15 |
| $\nu$ = 0.075 | 2/4 | 1.02 | 2.01 | 1.00 | 0.99 | 0.73 | 1.58 | 0.14 |
| $\nu$ = 0.30 | 4/4 | 1.05 | 0.50 | 1.00 | 1.00 | 0.73 | 1.57 | 0.14 |
| $\omega$ = 0.27 | 4/4 | 1.07 | 0.99 | 1.00 | 1.01 | 0.73 | 1.57 | 0.15 |
| $\omega$ = 1.08 | 2/4* | 1.37 | 199.7 | 0.86 | 0.01 | 0.59 | 2.74 | 0.11 |

Chains used: chains retained after excluding those that did not mix with the others; maximum $\hat{R}$ is for the retained chains (across all four chains it was 2.08 for $\nu$ = 0.075 and 1.99 for $\omega$ = 1.08). *No two chains converged for $\omega$ = 1.08; the two most consistent chains are shown for completeness only. FOI and reporting ratios are geometric means of the ratio of scenario to baseline posterior medians, across provinces (mean annual FOI) and health region-years (reporting proportion). Spearman (FOI): rank correlation of province mean annual FOI with baseline. $\rho$: relative susceptibility; CFR: case-fatality rate among reported cases.

### Supplementary figures

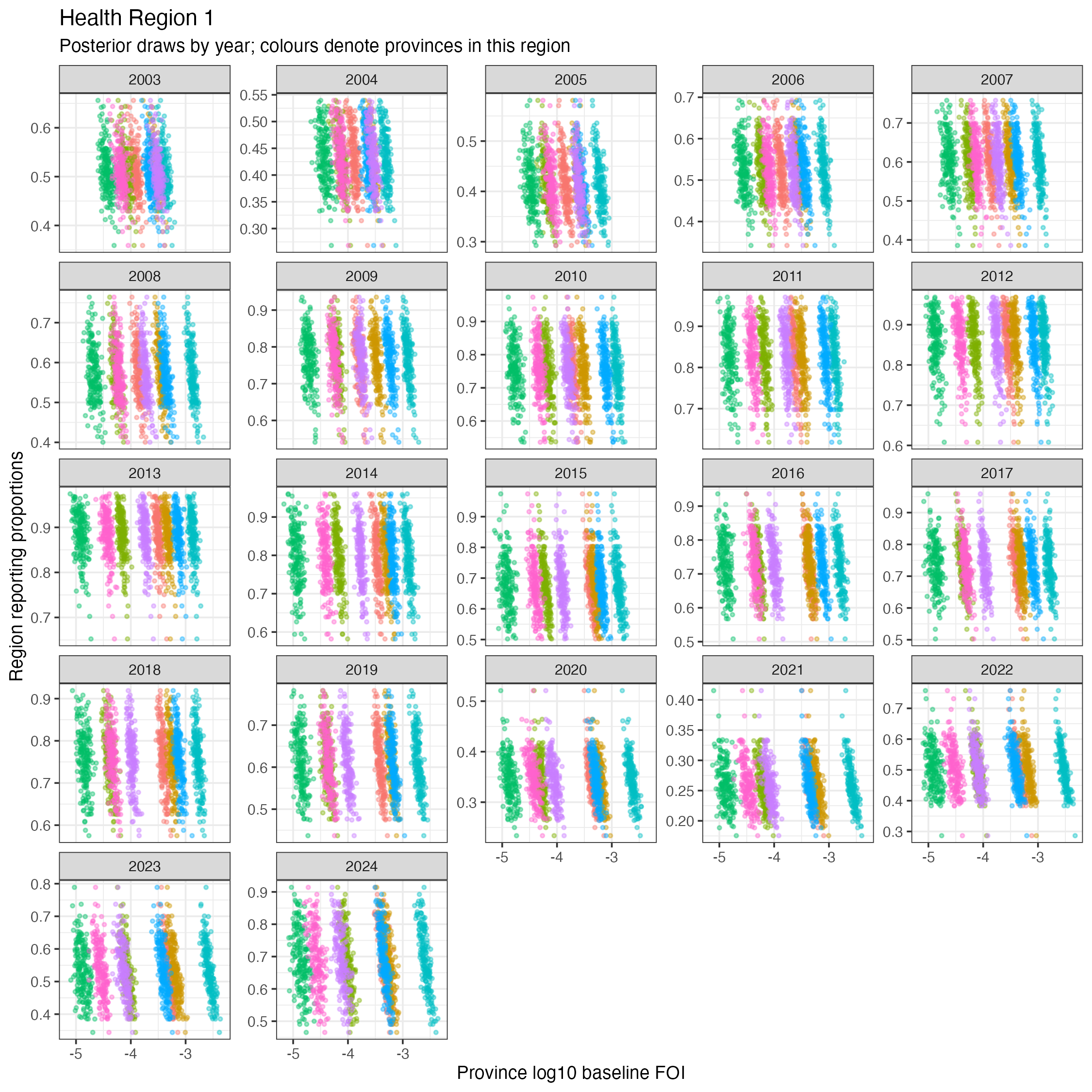

**Figure S1 Joint posterior distributions of baseline force of infection and reporting proportion.** Each panel shows posterior draws for one year; colours denote provinces within Health Region 1. The negative relationship within each province reflects the fact that reported cases depend on the product of transmission intensity and reporting proportion, so that a higher transmission intensity combined with lower reporting produces the same expected number of reported cases. Reporting proportion is estimated at the regional level and is therefore constrained jointly by all provinces in the region. Correlation plots across all regions can be found in Supplementary Files.

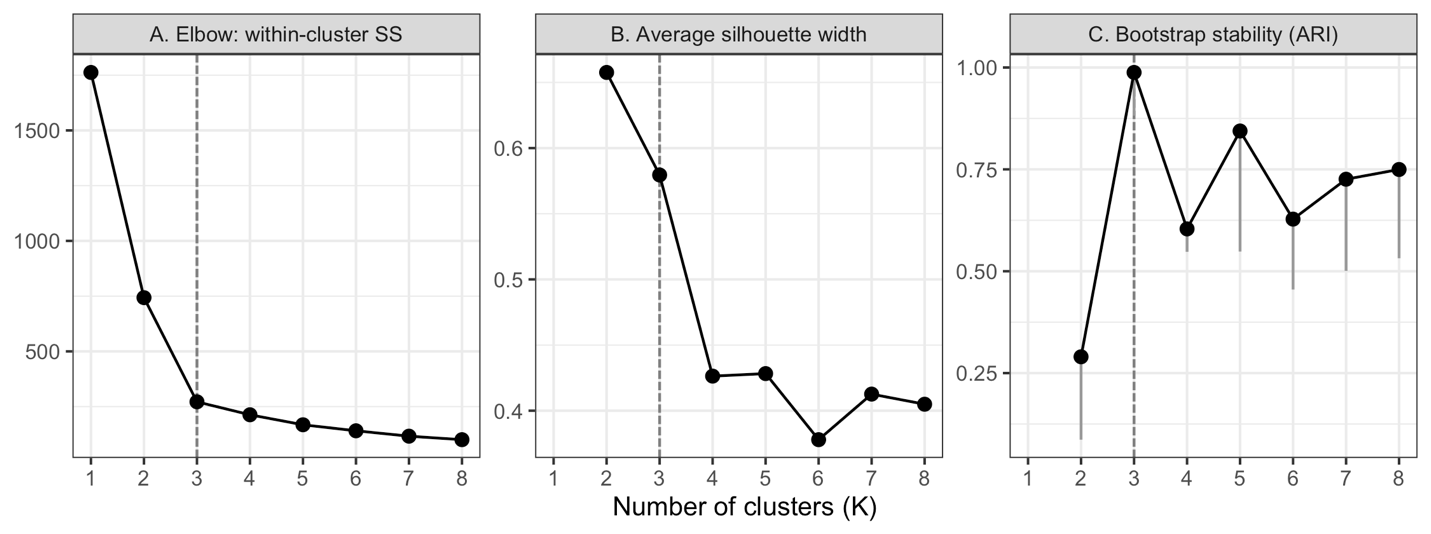

**Figure S2 Selection of the number of seasonal clusters.** (A) Within-cluster sum of squares (elbow method), (B) average silhouette width and (C) cluster stability, shown as the median adjusted Rand index between the full-data partition and partitions refitted on 200 bootstrap resamples (grey lines extend to the 5th percentile), for $K$ = 1–8. The dashed line marks the selected $K$ = 3.

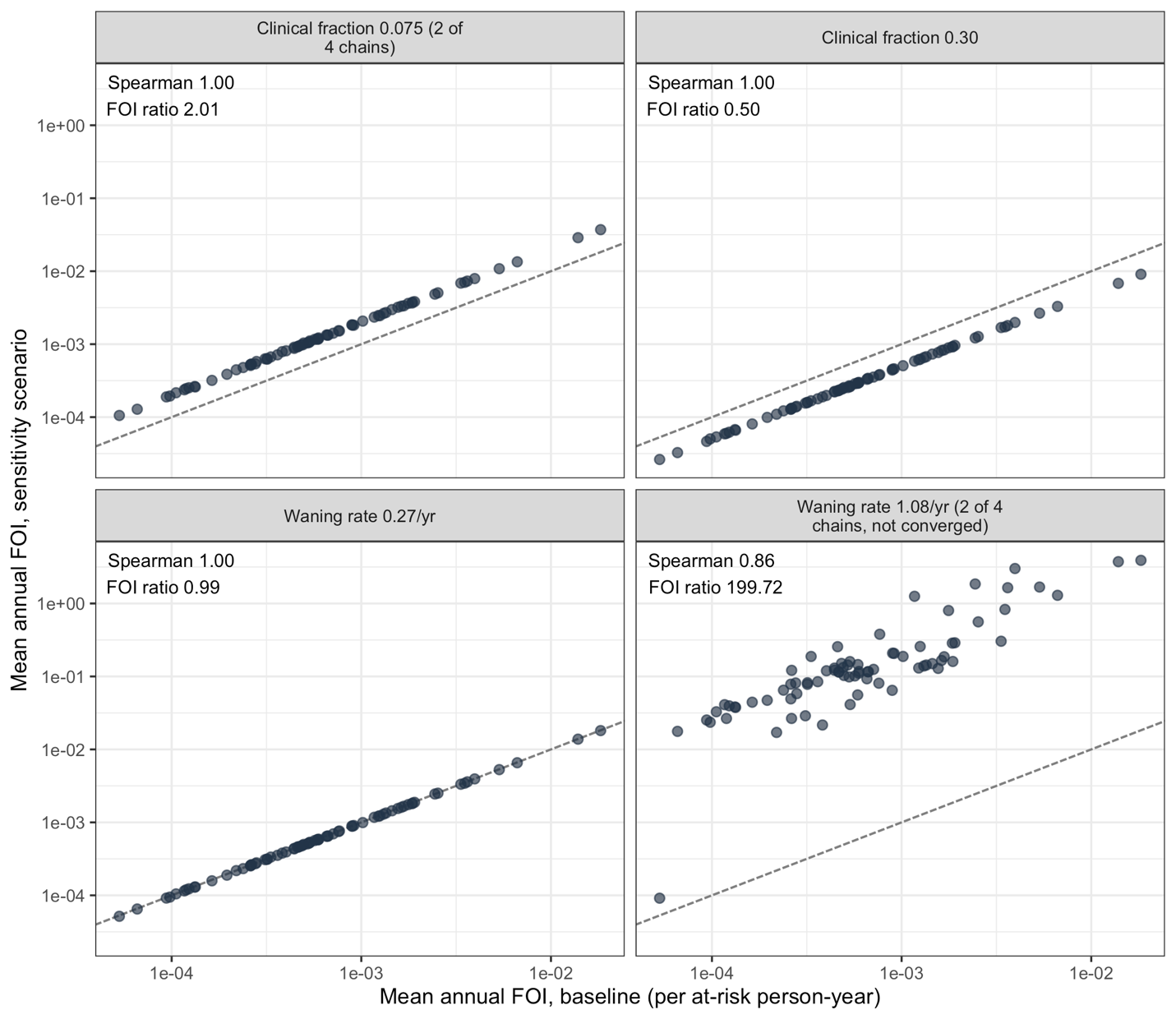

**Figure S3 Sensitivity of the estimated force of infection to the clinical fraction and IgG waning rate.** Each point is a province; axes show the mean annual force of infection over 2003–2024 (posterior median) under the baseline ($\nu$ = 0.15, $\omega$ = 0.54 ${year}^{-1}$) and each sensitivity scenario. The dashed line is the line of equality. Spearman rank correlations and geometric mean ratios (scenario/baseline) are shown in each panel. Where indicated, results are from the two of four chains retained; the scenario marked ‘not converged’ should not be interpreted quantitatively (Text S4).

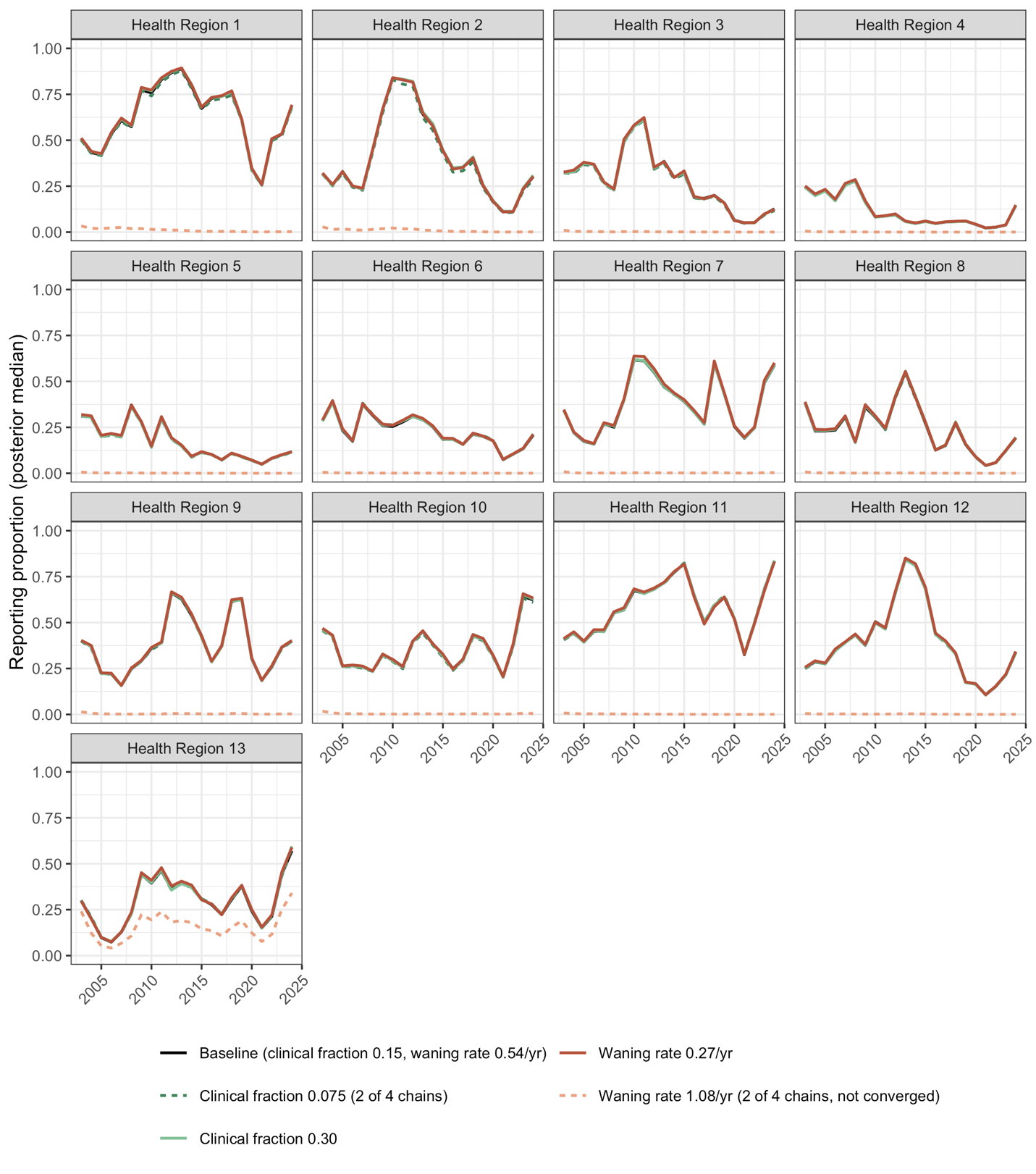

**Figure S4 Sensitivity of the estimated reporting proportion to the clinical fraction and IgG waning rate.** Lines show posterior medians by health region and year; dashed lines indicate scenarios summarised from a subset of chains (Text S4).

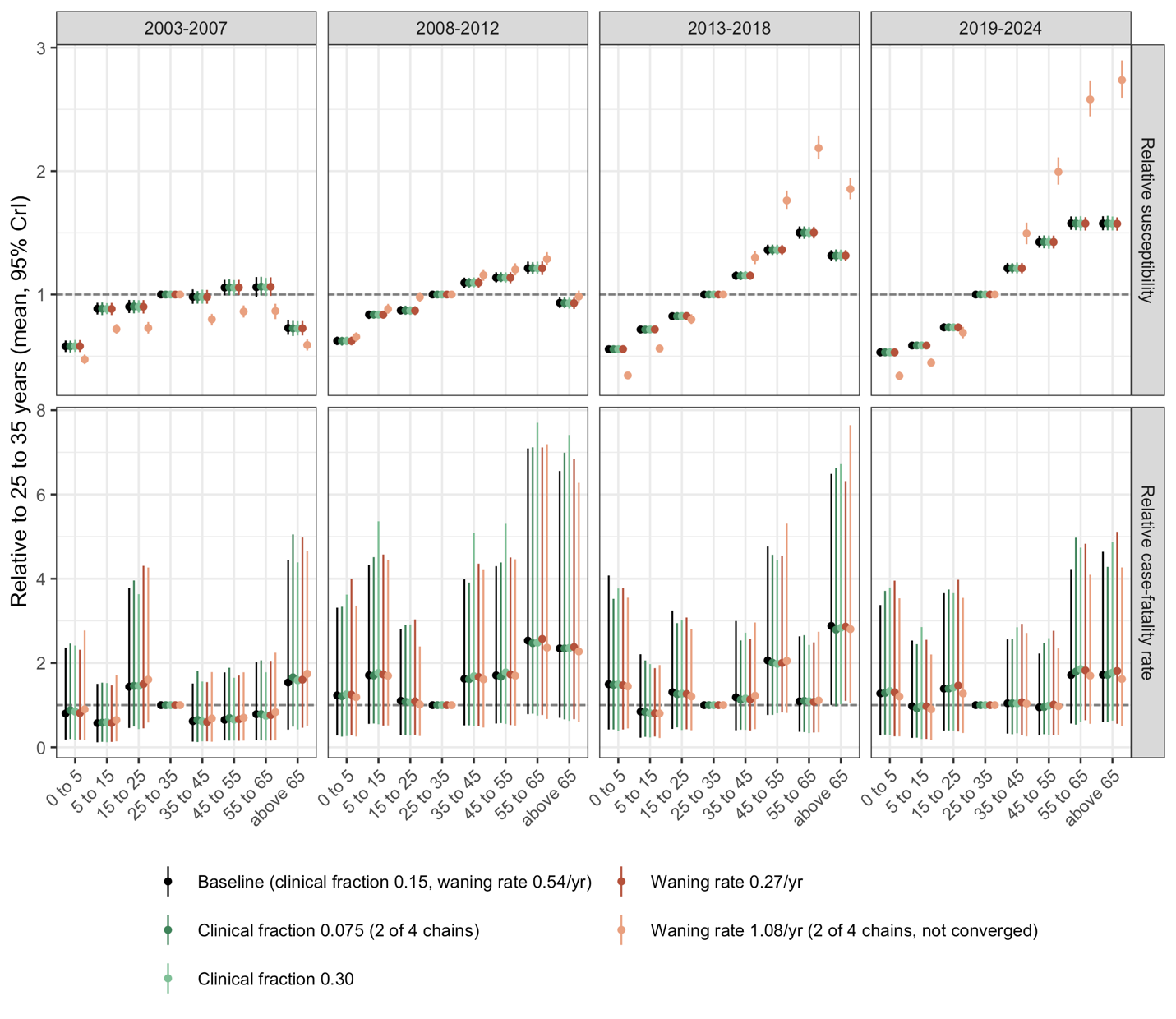

**Figure S5 Sensitivity of age-specific relative susceptibility and relative case-fatality rate to the clinical fraction and IgG waning rate.** Top: relative susceptibility; bottom: relative case-fatality rate among reported cases. Points and bars show posterior means and 95% credible intervals relative to the 25–35-year reference group. Where indicated, results are from the two of four chains retained; the scenario marked ‘not converged’ should not be interpreted quantitatively (Text S4).

### References

1.	Devamani C, Alexander N, Chandramohan D, Stenos J, Cameron M, Abhilash KPP, et al. Incidence of scrub typhus in rural south india. New England Journal of Medicine. 2025;392: 1089–1099. doi:[10.1056/NEJMoa2408645](https://doi.org/10.1056/NEJMoa2408645)

2.	Aiemjoy K, Katuwal N, Vaidya K, Shrestha S, Thapa M, Teunis P, et al. Estimating the seroincidence of scrub typhus using antibody dynamics after infection. Am J Trop Med Hyg. 2024;111: 267–276.

3.	Gonwong S, Mason CJ, Chuenchitra T, Khantapura P, Kijsri S, Tawan P, et al. Nationwide seroprevalence of scrub typhus, typhus, and spotted fever in young Thai men. Am J Trop Med Hyg. 2022;106: 1363–1369.
